# AI-Based Pipeline for the Segmentation of White Matter Hypoattenuations in CT Scans: A Design-Choice Validation Study

**DOI:** 10.64898/2026.03.10.26348006

**Authors:** Nada Alamoudi, Maria Del C. Valdés Hernández, Sohan Seth, Benjamin Jin, Eleni Sakka, Carmen Arteaga-Reyes, Daniela Jaime García, Yajun Cheng, Angela C.C. Jochems, Grant Mair, Joanna M. Wardlaw, Miguel O. Bernabeu

**Affiliations:** Institute for Adaptive and Neural Computation, Data Science and Artificial Intelligence, School of Informatics, University of Edinburgh, Edinburgh EH89AB, UK; Computer Science Department,The Applied College, Taibah University, Madinah 46537, Saudi Arabia; Institute for Neuroscience and Cardiovascular Research, Center for Clinical Brain Sciences, University of Edinburgh, Edinburgh EH16 4SB, UK; Department of Neurology, West China Hospital, Sichuan University, Chengdu 610041, China; Usher Institute, Center for Medical Informatics, University of Edinburgh, Edinburgh EH164UX, UK

**Keywords:** white matter hyperintensities, white matter hypoattenuations, Computed Tomography, Paired CT-MRI Data, Deep Learning, nnU-Net

## Abstract

**Purpose:** White matter hyperintensities are a key imaging marker of vascular pathology, defined on brain magnetic resonance imaging (MRI) and typically manifesting on non-contrast computed tomography (CT) as subtle white matter hypoattenuation (WMH). Accurately segmenting WMH in CT scans remains challenging due to their low contrast with the surrounding tissue. This work presents an end-to-end framework for WMH segmentation in CT scans and validates the design choices in each step of the processing pipeline. We leverage a state-of-the-art deep-learning method combined with manually annotated and pseudo-labelled datasets from paired CT-MRI scans from different clinical scanners to deliver reliable outcomes.

**Approach:** Our framework includes DICOM data curation, sequence selection, and automatic label generation as preparation steps. Preprocessing includes z-score intensity normalisation, skull stripping, CT windowing and two-step CT-MRI registration to accurately transfer MRI-derived labels into the CT space. Further processing involves the use of a 3D nnU-Net initially trained on CT images with aligned MRI-based WMH manually derived (n=91) and fine-tuned with two additional pseudo-labelled datasets (n=191).

**Findings:** CT-based WMH volumes showed a near-perfect correlation with ground-truth MRI WMH volumes (r = 0.98), with a systematic overestimation (mean difference = 2.40 mL; 95% limits of agreement: -8.31 to 13.11 mL) that may be adjustable in downstream tasks. This overestimation reflected challenges in the precise delineation of small WMH lesions and confounding from other imaging markers of brain disease. Across the evaluated cohort, ground-truth WMH volumes ranged from 1.02 to 149.34 mL. The best-performing configuration achieved a mean absolute error below 3 mL, corresponding to approximately 17% of the mean WMH volume, and a mean Dice similarity coefficient of 0.57. Segmentation accuracy decreased in the presence of stroke lesions. Models trained on single-pathology datasets, as well as approaches relying on template-based spatial normalisation, did not achieve satisfactory performance despite using the same backbone network configuration.

**Conclusion:** Using a multi-centre dataset and a multi-modal approach with expert-annotated data combined with pseudo-labelled data for training can substantially narrow the performance gap between CT- and MRI-based WMH segmentation. The framework proposed provides a generalisable solution that underscores the practical viability of CT for evaluating WMH burden in clinical and research scenarios—particularly where MRI is unavailable or contraindicated—thereby broadening access to small-vessel disease assessment.

## 1 Introduction

White matter hyperintensities (WMH) are regions of high signal intensity in T2-weighted and fluid attenuated inversion recovery (FLAIR) magnetic resonance imaging (MRI) brain scans, hypoattenuated (i.e., hypodense, less attenuation of the x-ray beam) in computed tomography (CT) scans. They are common in older adults and are a surrogate marker of small vessel disease (SVD). An increased burden of WMH has been strongly associated with cognitive decline, gait impairment, and with an increased risk of stroke, dementia, and mortality [1]. A high burden of WMH has been also associated with a higher likelihood of complications in ischaemic stroke patients after thrombolysis [2] and a worse prognosis after carotid endarterectomy [3].

Not surprisingly, a myriad of automated and semi-automated WMH segmentation algorithms have been developed [4] mostly for MRI, as it is the preferred imaging modality for WMH assessment [1, 5, 6]. However, CT is much more commonly used in clinical practice, especially in the emergency setting, such as acute stroke assessment [7]. CT is also preferred when MRI is contraindicated, for example, in patients with pacemakers or for patients uncomfortable with MRI scans that generally have longer acquisition times. Therefore, various automated methods to assess WMH directly from CT scans have been proposed.

Pitkänen et al. [8] and Kaipainen et al. [9] used convolutional neural networks (CNNs) to segment WMH from CT images. Pitkänen et al. [8] reported a moderate Dice Similarity Coefficient (DSC) of 0.43, which remains substantially lower compared to MRI-based methods where DSC values typically exceed 0.80 and often approach human-level performance (*DSC* ≈ 0.80) [10, 11] whilst Kaipanen et al. reported a correlation of 0.86 between computational measurements and visual clinical scores (i.e., Fazekas [12] scores). Chen and colleagues [13] segmented WMH in a multicenter cohort of patients with acute ischaemic stroke and obtained a correlation of 0.71 with expert-delineated segmentations, not statistically significantly different from the correlation between expert-delineated WMH masks and automatic WMH segmentations in the MRI-paired scans. Equally MRI-comparable results were obtained by Hanning and colleagues [14]. WMH segmentation results by van Voorst et al. [15] highly correlated with functional outcomes and with the appearance of symptomatic intracerebral hemorrhages, indicating that CT-based segmentation models can achieve clinically meaningful results. However, they obtained lower spatial overlap scores in external validations (DSC *<* 0.50), highlighting the potential and the need for further improvement in CT-based WMH segmentation accuracy.

In recent years, biomedical image segmentation tasks have increasingly benefitted from improvements and combinations of CNNs with encoder-decoder architectures, such as U-net [16]. As such, nnU-Net [17] has positioned itself as the state-of-the-art segmentation method. Ruy and colleagues [18] use it to segment WMH in a large (i.e., n=482/390 CT scans for training/validation) and heterogeneous sample, achieving a DSC of 0.527, with correlations of 0.832–0.891 with Fazekas visual scores consistent across CT vendors.

However, despite several methods being proposed, current CT-based WMH segmentation models still perform poorly in terms of spatial agreement with reference segmentations compared to MRI-based models [15], and, mainly due to the scarcity of high-quality manually labelled CT datasets for training, confidence in model performance is still low. As factors that may influence segmentation methods’ performance have not been analysed, avenues for improvement are, at most, speculative. Ryu and colleagues used automatically derived labels from MRI scans and recognise the high subjectivity and difficulties in the delineation of WMH in CT scans. CT datasets are typically noisier than MRI ones and have lower tissue contrast [5], making it hard to differentiate and segment WMH manually. Within the MRI sequences, WMH are easier to distinguish in FLAIR. In CT scans, the lesions are not as clearly defined (see Figure 1), leading to a lack of consistently labelled high-quality CT datasets.

**Figure 1:**
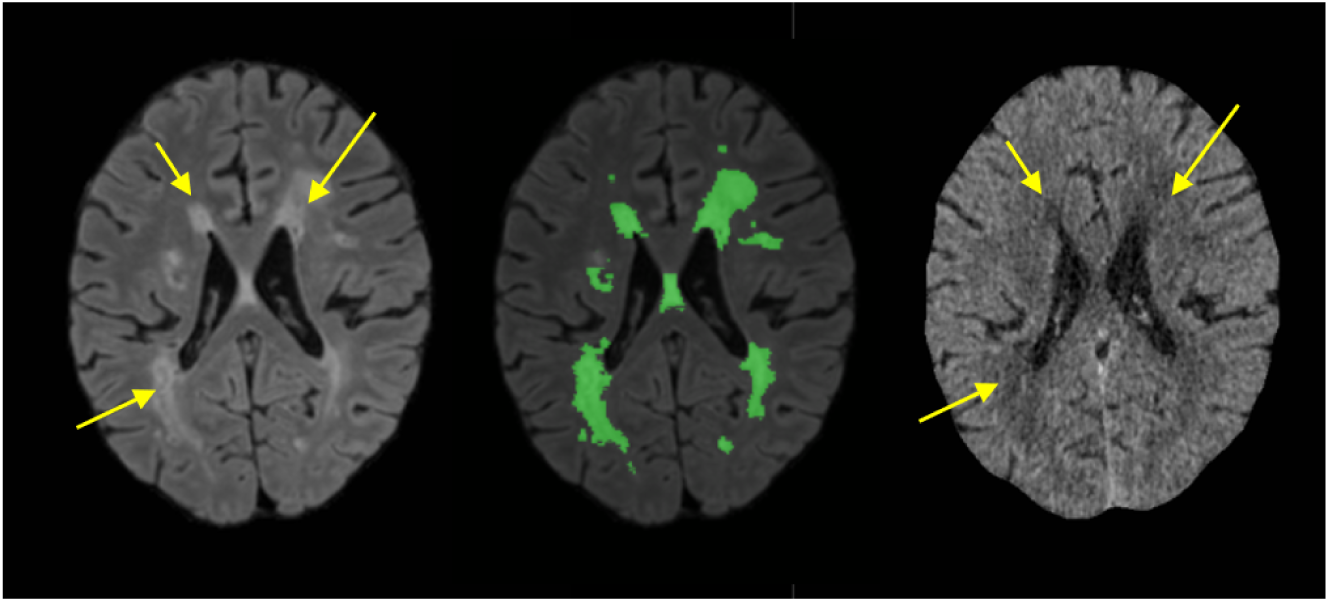
Representative axial brain images demonstrating the superior visibility of WMH, indicated by arrows, on MRI/FLAIR compared to CT. On the left: a raw MRI/FLAIR axial slice; in the centre: the same MRI/FLAIR slice with WMH automatic segmentations overlaid in green; and on the right: the co-registered CT image, where WMH are far less conspicuous. This contrast highlights the difficulty of reliably deriving high-quality training labels directly from CT data.

We address these limitations through an approach that combines manually labelled research data with pseudo-labelled (i.e., automatically annotated) clinical image data acquired at different scanners. By utilising pseudo-labelled clinical image data, we can increase the size of the training set and minimise the dependence on manually labelled data while allowing the model to generalise to real-world scenarios. We leverage diverse data sources with different white matter damage degrees and confounding factors to make the main model robust to different clinical settings. Through a careful analysis of each choice in the design of the steps of a full pipeline that includes clinical data curation, preprocessing, pseudolabelling, in addition to the actual feature segmentation, we seek to provide an end-to-end framework for CT-based WMH segmentation that achieves high performance and generalisability. As our backbone, we also use nnU-Net [17] to automate network design and hyperparameter optimisation. Our aims and contributions are four-fold: 1) to provide an end-to-end clinically applicable pipeline that bridges the gap between MRI- and CT-based WMH segmentation performance, 2) to improve the utility of routinely acquired non-contrast CT brain scans for WMH analysis to support clinical research and practice in situations where MRI may not be feasible or available, 3) to document and validate each design choice in our processing pipeline against previously published approaches to facilitate the interpretability of the methodological decisions and inform future optimisation, and 4) to identify the main factors driving segmentation performance and CT–MRI disagreement, including the detrimental effect of template-based spatial normalisation, the challenge of segmenting mild/low-burden WMH, modality-related differences in WMH volume estimates, confounding from enlarged perivascular spaces, and the impact of stroke lesions, whether chronic or acute, particularly when large.

## 2 Methods and Materials

### 2.1 Datasets

Three distinct datasets were used: the Mild Stroke Study 3 (MSS3) [19], the Third International Stroke Trial (IST-3) [20], and the CERMEP IDB MRXFDG (CIM) dataset [21]. These datasets span a wide variety of patient populations, scanning protocols, and imaging devices. As Figure 2 shows, we processed imaging data from 3,301 participants. Of them, 199 contributed to the development of the AI-based segmentation model (i.e., green blocks in Figure 2) and 229 (i.e., participants from MSS3) to the evaluation of the model and factors influencing its performance (i.e., fourth of the contributions listed above).

**Figure 2:**
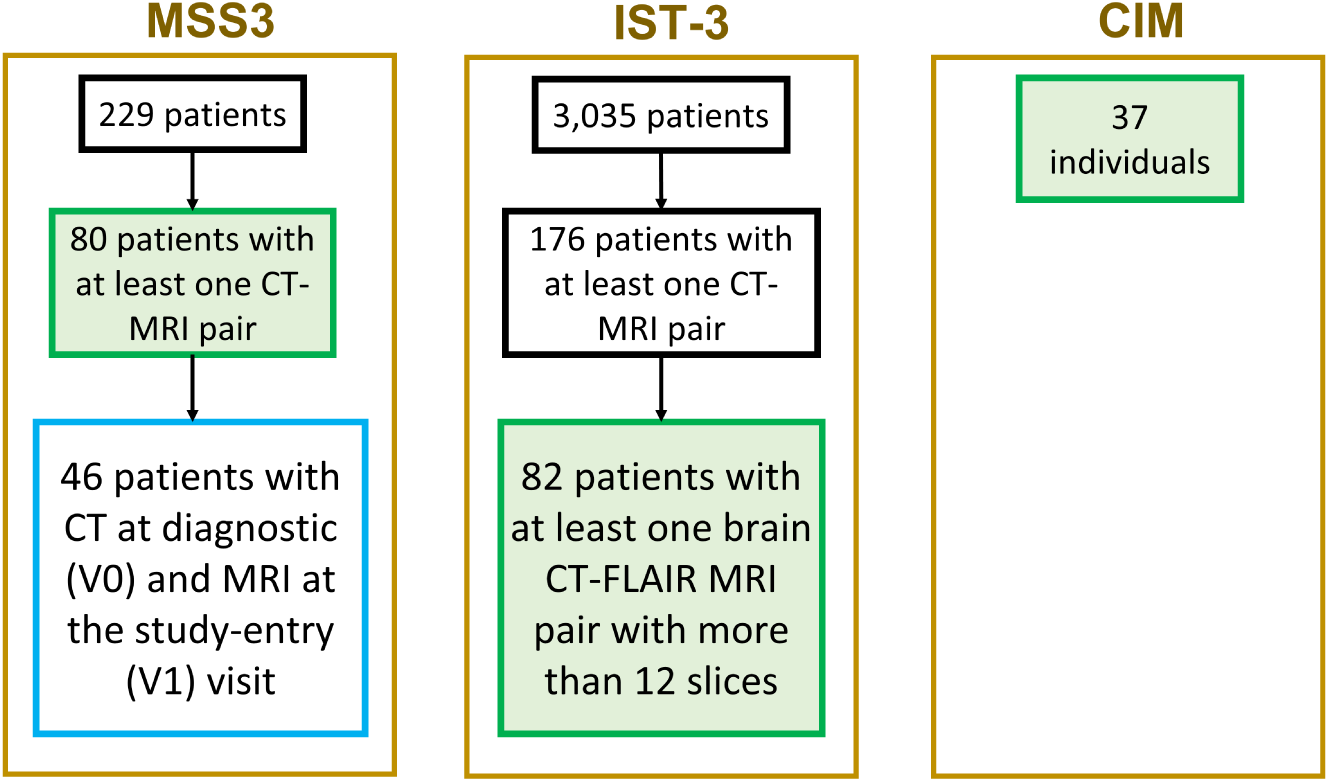
Number of patients processed from each contributing study database. In green blocks the number of study participants that contributed to the training and validation, and in the white block with blue border the number of study participants that contributed to the testing and explainability analyses.

#### 2.1.1 Mild Stroke Study 3 Dataset

The MSS3 is a prospective observational study focused on studying the evolution of small vessel disease (SVD) in patients who had a mild ischemic stroke (modified Rankin Scale ≤ 2) and presented to Edinburgh/Lothian stroke services [19] within hours of the onset of the stroke symptoms. The MSS3 enrolled 229 participants (≥ 18 years old) without major neurological or severe cardiorespiratory conditions, who provided informed consent to participate in the study. After the first clinical brain scan (that is, MRI or CT, in one of the four different hospitals in the region), study participants were invited to the Royal Infirmary of Edinburgh to undergo subsequent high-resolution 3T MRI scans with a research protocol. In the case of any recurrent symptomatic event, additional clinical brain scans (that is, MRI or CT) were also performed at one of the regional hospitals or at the same 3T research scanner. Ethical approval was obtained from the Southeast Scotland Regional Ethics Committee (18/SS/0044).

For our analyses we selected 91 MRI/FLAIR scans paired with the CT scans from 80 participants, taking care that 1) CT and MRI were acquired within approximately 3 months or less from each other, 2) the pairs were unique (i.e., each image belongs only to one pair), and 3) the MRI scans were acquired with the research protocol. The mean interval between CT and MRI images in the pairs selected was ∼ 6.68 days; the median interval was 37 days (IQR 55 days; 95% CI 28–44 days). The mean (SD) age of the patients that provided data for this study is 67.3 (11.3) years old. From them, 26 (32%) were female and 54 (68%) were male. The median WMH volume in this sample, measured in FLAIR, is 10.27 ml (IQR 5.10 to 22.34 ml).

To explore the influence of a) the time difference between CT and MRI scans and b) the differences between the two imaging modalities in the results, we additionally used the image data of the rest of the MRI scans acquired at the two time points that provided most of the CT-MRI image pairs: i.e., the diagnostic scan visit, which we subsequently denote as V0, and the study-entry visit, denoted as V1.

#### 2.1.2 Third International Stroke Trial Dataset

The IST-3 dataset derives from a large, international, multi-centre, randomised controlled trial to determine if intravenous recombinant tissue plasminogen activator (rt-PA) is effective in acute ischaemic stroke [20]. Ethical approval was obtained from the Multicentre Research Ethics Committees, Scotland (MREC/99/0/78) and local ethics committees. A total of 3,035 patients from 156 hospitals in 12 countries were enrolled in the trial.

Of this cohort, 176 patients were initially identified with both CT and MRI scans. From the 106 that had CT head scans with full or almost full brain coverage and acquired with tissue kernel, twenty-four patients were excluded because they had only sagittal or coronal FLAIR scans with too few slices—when reoriented to the axial plane these volumes were too low-resolution for reliable analysis. Only 82 had paired CT/FLAIR sequences and met our inclusion and quality-control criteria (data cleaning and selection process summarised in Figure 3) and formed our final IST-3 cohort. The average interval between CT and FLAIR MRI was 3.1 ± 4.9 days; the median interval was 2 days (IQR 3 days; 95% CI 1–2 days) (Supplementary Figure 1). This dataset primarily reflects acute ischaemic stroke images. The mean (SD) age of the 82 selected patients is 78.7 (10.7) years; of these, 38 (46.3%) were female and 44 (53.7%) were male. The median WMH volume in this sample, measured in FLAIR is 15.33 ml (IQR 4.88 to 34.13 ml)

**Figure 3:**
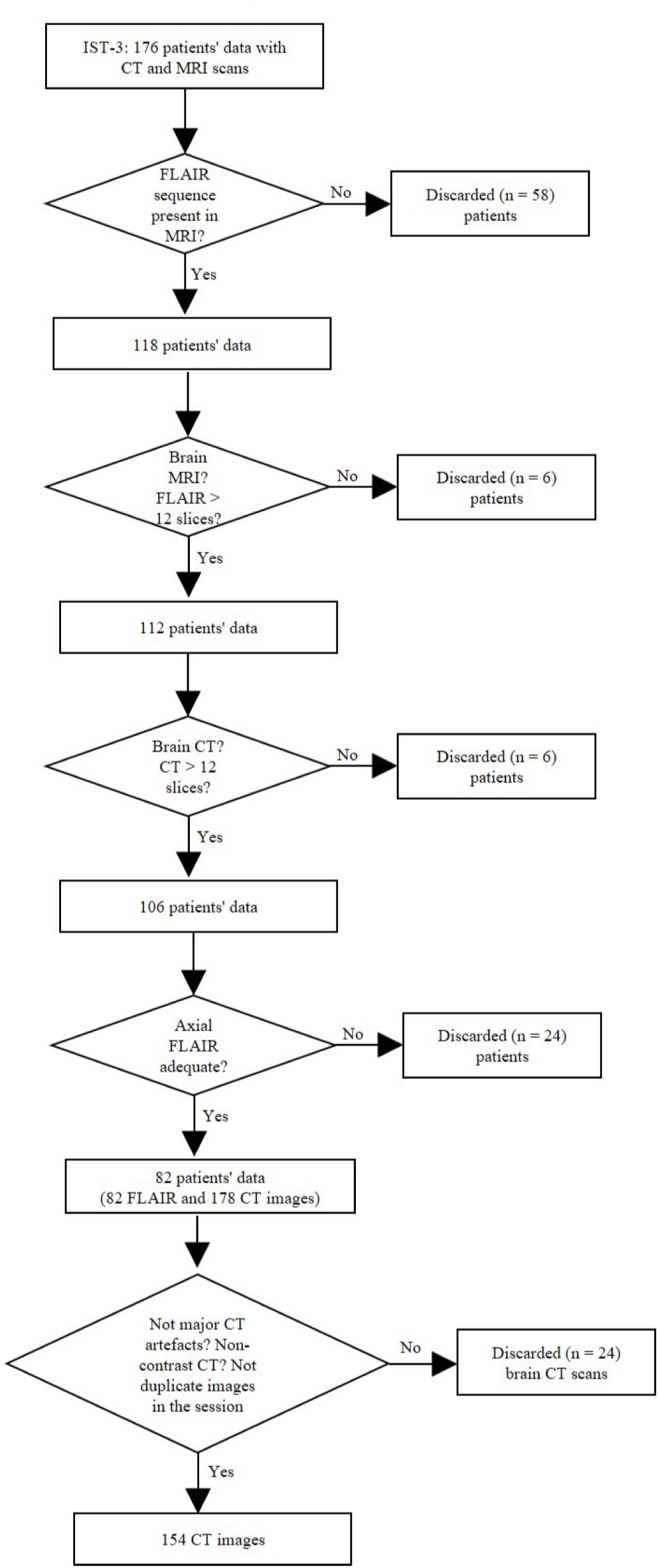
Flowchart of the patient selection and quality-control process for the Third International Stroke Trial (IST-3) dataset.

#### 2.1.3 CERMEP IDB MRXFDG Dataset

The CIM dataset (CERMEP IDB MRXFDG) comprises multimodal neuroimaging data from 37 healthy young adults [21] of mean (SD) age of 38.1 (11.4) years old, 54% of who were female. From this sample, only three individuals (8.1 %) presented mild WMH with median volume measured in FLAIR of 1.09 ml (IQR 0.72 to 1.70 ml). All imaging, including MRI/FLAIR and CT, was acquired on the same day. The CIM dataset provides a unique control population and an opportunity to refine the model’s ability to detect small and subtle WMH lesions and identify the absence of it.

### 2.2 Imaging Protocols

Across the MSS3, IST-3, and CIM datasets, imaging parameters (voxel size, slice thickness, scanner vendor, and field strength) differed substantially. The MR images used from the MSS3 were acquired at 3T with a high-resolution research imaging protocol. Both CT and MR images from the IST-3 had lower spatial resolution than those from the MSS3 dataset and were acquired using a variety of clinical protocols. The majority of MRI scanners that provided images for the present study had a magnet strength of 1.5 T. The CIM dataset used 1.5 T MRI scanners, but the imaging protocol involved FLAIR images with isotropic voxels of 1.2 mm^3^ resolution, with high image quality. A detailed summary of the key imaging parameters (voxel size, matrix size, slice thickness, MRI field strength, and scanner manufacturer) is shown in Table I. By incorporating this variability, we aim to improve the generalisability of the segmentation model trained on these datasets.

**Table I:** Summary of Imaging Parameters for Paired CT and FLAIR Scans Across Datasets

| Parameters | MSS3 Dataset |  | IST3 Dataset |  | CIM Dataset |  |
| --- | --- | --- | --- | --- | --- | --- |
|  | CT | FLAIR MRI | CT | FLAIR MRI | CT | FLAIR MRI |
| **Acquisition** | 3D | 3D | 3D | 3D | 3D | 3D |
| **MRI Field Strength** | N/A | 3T (100%) | N/A | 3T 10 (12.19%)<br>1.5T 72 (87.8%) | N/A | 1.5T (100%) |
| **Slice Thickness Range(mm)** | 0.2 – 5 mm | 0.9 mm | 0.5 – 9 mm | 4 – 7.8 mm | 1.5 mm | 1.20 mm |
| **< 5 mm** | 94.47% | 100.0% | 80.33% | 22.0% | 100% | 100% |
| **≥5 mm** | 5.52% |  | 19.66% | 78.0% |  |  |
| **Slices Number Range Minimum – Maximum** | 27 - 601 | 176 | 12 - 287 | 19 - 39 | 233 | 256 |
| **Voxel size (mm<sup>3</sup>)** | 0.4 × (0.4- 0.49) × (0.2 – 5) | 0.9 × 0.9 × 0.9 | (0.3 – 0.57) × (0.3 – 0.57) × (0.5 – 9) | (0.4 – 0.97) × (0.4 – 0.97) × (4 – 7.80) | 0.6 × 0.6 × 1.5 | 1.2 × 1.2 × 1.2 |
| **Matrix Size** | 512 × 512 × (27- 601) | 256 × 256 × 176 | 512 × (512- 697) × (12 – 287) | (196 – 512) × (256 – 512) × (19 – 39) | 512 × 512 × 233 | 176 × 196 × 256 |
| **Orientation** | Axial (100%) | Axial (100%) | Axial (100%) | Axial (77.4%)<br>Coronal/Sagittal (22.6%) | Axial (100%) | Axial (100%) |
| **Manufacturers** | Canon (55.67%)<br>Toshiba (16.75%)<br>Siemens (13.81%)<br>GE (7.91%) | Siemens Prisma (100%) | Philips (41.01%)<br>Siemens ( 21.35%)<br>GE (35.39%)<br>Toshiba (2.25%) | Philips (54.88%)<br>Siemens (24.39%)<br>GE (18.29%)<br>Unknown (2.44%) | Siemens Biograph mCT64 (100%) | Siemens Sonata (100%) |
| **Number of scanner models by manufacturer** | **N = 7**<br>▪ **Siemens:** SOMATOM Definition AS+, Definition Edge, Definition AS<br>▪ **Canon:** Aquilion ONE, Aquilion Prime SP<br>▪ **GE:** Optima CT660<br>▪ **Toshiba:** Aquilion | **N = 1** | **N = 18**<br>▪ **Philips:** Brilliance 16P, Brilliance 40, Brilliance 16, Brilliance16, Philips CT Secura<br>▪ **GE:** HiSpeed CT/i, LightSpeed QX/i, HiSpeed NX/i, LightSpeed Ultra, LightSpeed16, LightSpeed VCT, BrightSpeed, LightSpeed Pro 32<br>▪ **Siemens:** Sensation 16, Sensation 64, Definition AS+, Definition AS<br>▪ **Toshiba:** Aquilion | **N = 9**<br>▪ **Siemens:** Avanto, Symphony, Verio<br>▪ **Philips:** Inters, Achieva<br>▪ **Unknown:** Eclipse 1.5T<br>▪ **GE:** SIGNA HDx, GENESIS SIGNA, Signa HDxt | **N = 1** | **N = 1** |

### 2.3 Image Preprocessing and Registration

#### 2.3.1 Data Preparation

With the exception of the MSS3 MRI data, which were available in the Neuroimaging Informatics Technology Initiative (NIfTI) format, the rest were in the Digital Imaging and Communications in Medicine (DICOM) format. We automatically processed the DICOM tags to identify and remove localisers, images with less than 12 slices, angiograms, dynamic MRI sequences, contrast-enhanced images, and CT images with bone kernels. From MRI DICOM tags we also extracted the flip angle, echo time (TE), repetition time (TR), and inversion time (TI) parameters to further identify the sequences. We identified the MRI sequences in two complementary ways: 1) querying the tags reserved for the sequence name (i.e., series description (tag 0008,103E), sequence name (tag 0018,0024), scanning sequence (tag 0018,0020), and sequence variant (tag 0018,0021)) and using an in-house lookup table with more than 100 possible names, and 2) applying a rule-based decision tree classifier that uses the parameters of the MRI sequence to estimate the type and assign a uniform name (e.g., T1-weighted, T2-weighted, FLAIR). Table II shows the range of each parameter considered for each sequence type. The complementary strategy for sequence identification was necessary due to the heterogeneity and sometimes absence of information in relevant DICOM tags.

**Table II:** MRI sequence parameters including repetition time (TR), echo time (TE), flip angle (FA), and inversion time (TI), used for MRI sequence classification from DICOM tags. These parameters guided the assignment of standardised sequence names during preprocessing. Abbreviations: T1W (T1-weighted), PD (Proton Density-weighted), T2W (T2-weighted), FSE (Fast Spin Echo), GRE (Gradient Echo), STIR (Short Tau Inversion Recovery), FLAIR (Fluid Attenuated Inversion Recovery).

| Sequence | Repetition Time<br>(TR, ms) | Echo Time<br>(TE, ms) | Flip Angle<br>(FL, °) | Inversion Time<br>(TI, ms) |
| --- | --- | --- | --- | --- |
| T1W | <800 | <30 | 70–110 | - |
| PD | >1000 | <30 | 90 | - |
| T2W | >2000 | >80 | 90 | - |
| GRE T2*W | Variable | <30 | 5–20 | - |
| STIR | >2000 | >60 | 180, 90 | 120–170 |
| FLAIR | >3000 | >80 | Variable | 1700–2200 |

#### 2.3.2 Format Conversion and Quality Control

All image data were converted from DICOM format to NIfTI format using dcm2niix [22] (https://github.com/rordenlab/dcm2niix) and organised as per the Brain Imaging Data Structure (BIDS) specifications (https://bids.neuroimaging.io/index.html). dcm2niix is a robust and popular tool due to its high reliability across most protocols. To adhere to the NIfTI format, dcm2niix resamples CT imaging to equidistant slices to remove possible gantry tilt. However, dcm2niix also sometimes resamples due to floating-point errors in the calculation of the slice distances. Supplementary Figure 2 shows the histogram of the intensity values of all matched DICOM and NIfTI slices within the brain window in an example CT image series from the IST-3 database, illustrating the impact of this unnecessary resampling. While this effect is most prominent at bone edges, it is also observable in soft tissue. An influence on segmentation masks can therefore not be ruled out.

Further visual inspection identified incomplete or unusable data, such as images with limited brain coverage (e.g., base or top of the brain only) or those with severe motion artefacts, which were also excluded.

#### 2.3.3 Intensity Normalisation and Skull Stripping

CT intensities were clipped to the typical Hounsfield Unit (HU) range of [-1024, 3071] [23], and a standardised “brain window” (Window Level = 40, Window Width = 80) was applied to enhance brain parenchyma visibility. From the MRI sequences, we used FLAIR for our task, as it shows the WMH with higher conspicuity. Both CT and MRI/FLAIR scans underwent skull stripping using SynthStrip [24] (https://surfer.nmr.mgh.harvard.edu/docs/synthstrip/), a deep learning–based tool from Harvard University to remove non-brain tissues.

#### 2.3.4 Registration Procedures

Linear registration methods aligned the FLAIR-based WMH masks with the CT anatomy. FSL FLIRT [25, 26] and NiftyReg (//cmictig.cs.ucl.ac.uk/wiki/index.php/NiftyReg) tools were evaluated to determine the optimal alignment strategy. The chosen approach involved rigid-body followed by affine linear transformations, enabling the transferring of WMH masks from the MRI space to the CT domain with minimal anatomical distortion. These sequential steps ensured that the training targets accurately reflected WMH lesions in the CT space, consequently improving subsequent WMH segmentation performance. Non-linear registration was considered, but not chosen for two reasons. Firstly, the DICOM to NIfTI conversion already introduced alterations in the images while homogeneising the slice thickness. We want to minimise the number of spatial transformations that could be a source of alterations and distortion. Secondly, to validate the results from the segmentation *per se* we need to transform the resultant WMH masks from the CT space into the MRI space where we have our reference segmentations. Only linear space transformations guarantee reversibility. Despite claims that diffeomorphic registration is reversible, especially when using a symmetric block-matching approach like the one used by niftyreg, authors of the method reported a mean error of 1.64 (maximum error of 3.33) when transforming from a T2-weighted MRI space to CT [27]. We also investigated the impact of aligning all CT images to a template (e.g., to a CT or MRI template [28]) prior to input into the segmentation framework, as a form of spatial normalisation suggested by previous works [8, 9, 13].

#### 2.3.5 Data Integration pipeline

As illustrated in the pipeline diagram (Figure 4), we combined three datasets: MSS3 (n = 91 - from 80 patients - with expert WMH masks), IST-3 (n = 154 - from 82 patients - with pseudo-labels) and CIM (n = 37 with pseudo-labels) to cover a broad spectrum of image qualities, scanner types, and clinical contexts. This approach aimed to increase model generalisability and decrease dependence on a single, high-quality dataset. With high-fidelity manual annotations from MSS3 data and additional training with pseudo-labelled IST-3 and CIM data, the model could learn to identify robust WMH representations from mild to severe lesion burdens accompanied by other imaging markers of vascular pathologies in different degrees or the absence of them across patient populations.

**Figure 4:**
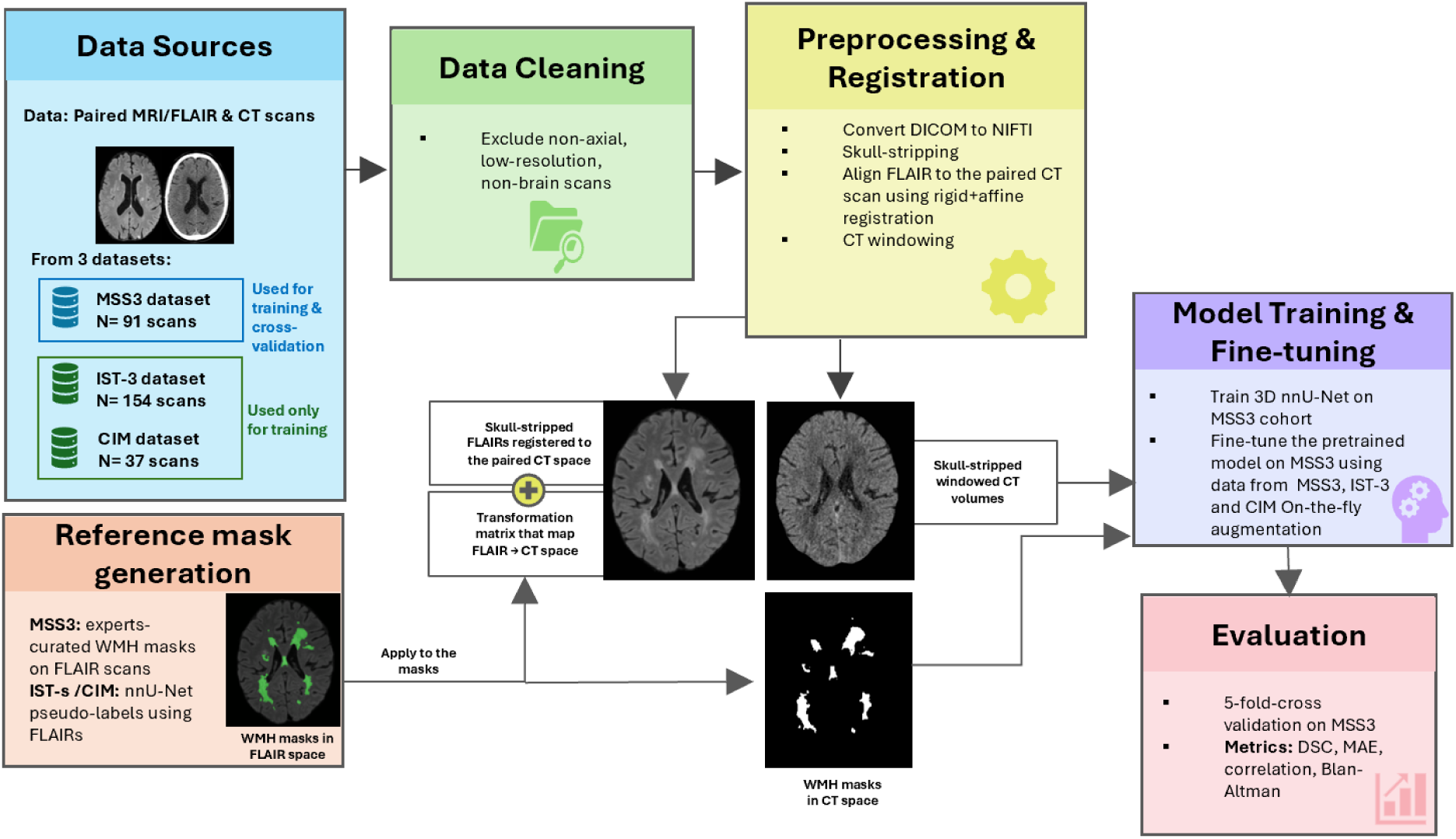
Graphical Abstract of our end-to-end CT-based WMH segmentation pipeline.

### 2.4 Generation of Reference WMH Masks

We used MRI/FLAIR as the modality for generating reference WMH segmentations, which has superior WMH-background contrast than CT and well-established protocols for WMH delineation [5,6] to establish robust training targets.

For the MSS3 dataset, WMH masks were generated semi-automatically by a team of three experienced image analysts to provide high-fidelity ground truth labels. These annotations were created following standardised methods designed to assess SVD markers, including WMH, stroke lesion volumes, and related neuroanatomical metrics [19, 29]. Briefly, the intensities of the brain parenchyma were standardised. Intense WMH priors were considered those voxels with values above 1.7 times the z-scores of the brain tissue intensities, and less intense WMH priors were those voxels with values above 1.3 times the z-scores of the brain tissue intensities. These initial priors were subsequently refined using brain ventricles, aqueduct, choroid plexus and cisterna magna masks generated from FreeSurfer (https://surfer.nmr.mgh.harvard.edu/), a probability WMH distribution map generated from 600+ segmentations from healthy ageing individuals, and a hierarchical filtering using a Gaussian kernel. The results were inspected for accuracy, and false positives, as well as ischaemic stroke lesions, were manually edited out from the masks. This subset of data, with “gold-standard” manual WMH annotation, served as the foundation for model training and validation.

For the IST-3 and CIM datasets, we generated WMH labels automatically using a previously validated, pre-trained 3D nnU-Net model applied to MRI/FLAIR scans [30]. This “pseudo-labelling” approach allowed us to expand the size and diversity of the training dataset without the labour-intensive manual annotation. The FLAIR-based WMH masks were subsequently mapped into the corresponding CT space using the MRI-to-CT transformations previously explained.

### 2.5 Deep Learning Model: nnU-Net Framework

For segmenting WMH in the CT scans we employed nnU-Net [17], a state-of-the-art deep learning framework that was previously applied to this task [15], [18] yielding the best performance among the reported methods applied up to date. This out-of-the-box tool trains two basic types of networks: 2D U-Net and 3D U-Net to perform semantic segmentation of 3D images with high accuracy and performance. It systematises the configuration process on a task-agnostic level to drastically reduce the need for tuning the network automatically configuring the architectures and hyperparameters to the characteristics of a given input dataset, thus simplifying network architecture optimisation and reducing the need for extensive manual tuning.

However, different from the work from Ryu et al. [18], who used a 2D configuration, our optimal configuration was a 3D residual encoder U-Net [16], arranged in a six-stage encoder-decoder architecture. Each encoder stage successively increased the number of feature channels (from 32 up to 320) and integrated multiple residual blocks with 3D convolutions (kernel size [3, 3, 3]). Instance normalisation and LeakyReLU activations were applied at every layer, stabilising training and ensuring efficient gradient flow. The input volumes were resampled to approximately 1.5 × 0.488 × 0.488 mm^3^ per voxel, and the network was trained on patches of size 56 × 192 × 160 voxels with a batch size of two.

### 2.6 Model Training and Validation

Previous supervised learning models either use manually derived reference segmentations [15] or pseudo-labelled data [8], [9], [18] for training the machine-learning segmentation algorithms. We conducted three experiments using different training sets to investigate the impact of using pseudo-labelled data on the overall model performance: 1) MSS3 only, 2) MSS3 + CIM, and 3) MSS3 + CIM + IST-3. But we only used the MSS3 dataset to validate the models, as this was the only dataset from which we had expert-generated manually labelled data. We used five-fold cross-validation in all the experiments. In each fold we used for training the MSS3 cases not involved in the validation set. In the validation set, we split the MSS3 data taking care that in each fold were present different degrees of WMH loads. The distribution of WMH loads (i.e., classed in low, medium, high) across the five folds is shown in Figure 5 and Table III.

**Figure 5:**
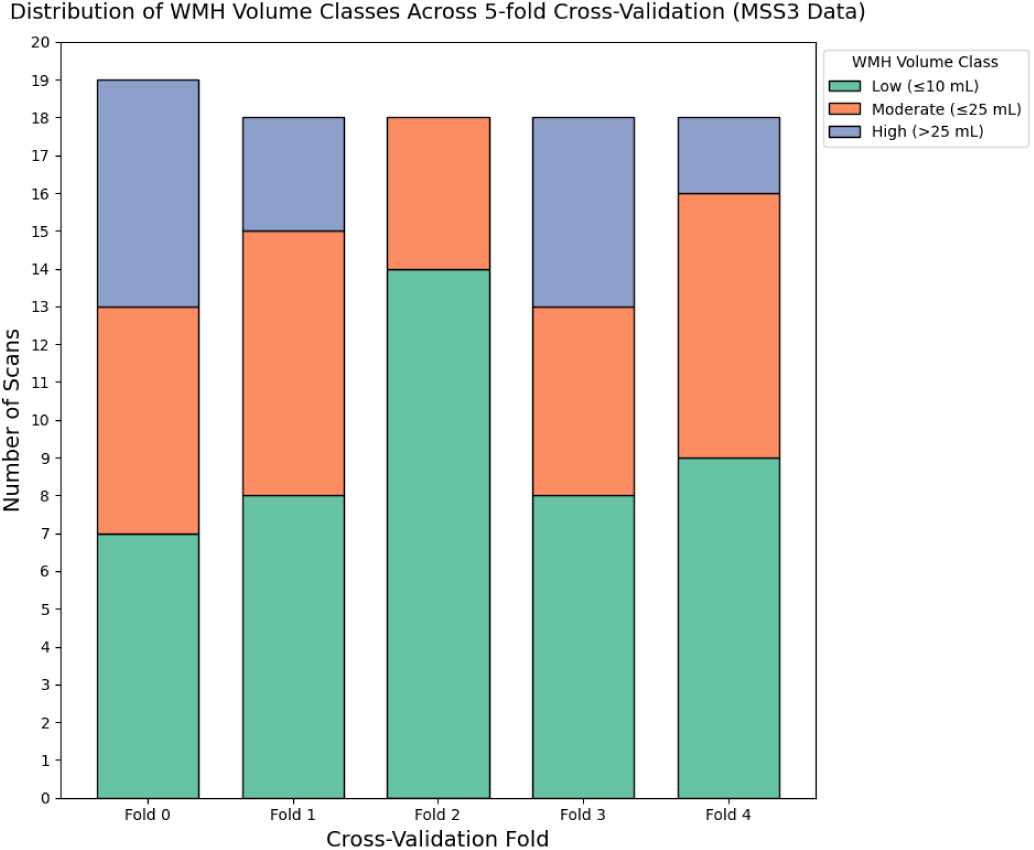
Distribution of WMH burden categories (low ≤10 mL, medium ≤25 mL, high *>*25 mL) across the five cross-validation folds of the MSS3 dataset. Each stacked bar represents a fold and illustrates the number of scans classified into each WMH burden category.

**Table III:** The number of scans for each WMH burden class in the MSS3 dataset.

| WMH Burden Category | N (Scans) |
| --- | --- |
| High Volume ($> 25$ mL) | 16 |
| Low Volume ($\leq 10$ mL) | 46 |
| Medium Volume ($\leq 25$ mL) | 29 |

We trained the network for a maximum of 1000 epochs, stopping early (criterion: 50 epochs without improving validation loss). This strategy allowed for efficient training with respect to the training parameters and, at the same time, mitigated the risk of overfitting. To address the class imbalance and segmentation accuracy, we used a default stochastic gradient descent optimiser and a composite Dice–cross-entropy loss function [17].

### 2.7 Data Augmentation and Fine-Tuning

Additional model generalisability and mitigation of overfitting were achieved using on-the-fly data augmentation (e.g., rotations, scaling, and flipping) provided by nnU-Net. We also fine-tuned the model with pseudo-labelled IST-3 and CIM data to increase the diversity of lesion appearance and scanner parameters, and, hence, the model’s training distribution. The final performance was evaluated only on the MSS3 dataset to guarantee a consistent and reliable evaluation.

### 2.8 Evaluation Metrics

Segmentation performance was quantified using a comprehensive set of evaluation metrics, including spatial overlap measures, volumetric comparisons, and bias assessments.

Below are detailed definitions of the evaluation metrics used:

#### 2.8.1 Spatial Overlap and Detection Metrics

##### Dice Similarity Coefficient (DSC)

The DSC measures the spatial agreement between the predicted segmentation *S* and the ground truth *G*:

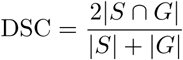

A DSC of 1 indicates perfect overlap, while 0 indicates no overlap.

##### Sensitivity (Recall)

Sensitivity is the model’s ability to correctly identify WMH regions:

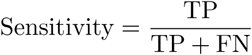

##### Precision (Positive Predictive Value)

Precision indicates how many positively predicted voxels are actually WMH:

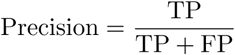

##### Regional false positive burden (FP burden)

for a region-of-interest mask *R* (basal ganglia (BG) and centrum semiovale (CSO)), as defined in [31], we compute the region-restricted false positive voxel count:

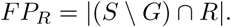

To normalise for ROI size, we report the regional false positive burden per mL of ROI (voxels/mL):

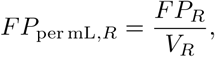

where *V_R_*is the ROI volume in mL.

##### Regional false positive concentration (FP concentration)

to quantify the spatial distribution of false positives relative to the total false positives in the volume, we compute

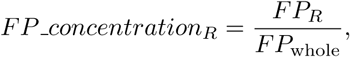

where *FP*_whole_ is the false positive voxel count computed over the whole evaluation domain (skull-stripped volume).

#### 2.8.2 Volumetric Agreement

##### Mean Absolute Error (MAE)

MAE quantifies the average magnitude of errors between predicted volumes *y*^*_i_* and ground truth volumes *y_i_* over *n* samples:

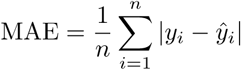

##### Pearson’s Correlation Coefficient (r)

Pearson’s *r* evaluates the linear relationship between predicted and ground truth volumes:

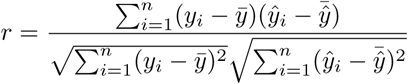

##### Bland–Altman Analysis

This analysis assesses the agreement between two measurement techniques by evaluating the mean difference (bias) and the limits of agreement. Let *y*^*_i_* be the predicted volume and *y_i_* be the ground truth volume:

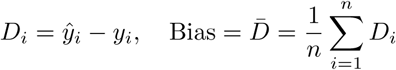

The 95% limits of agreement are given by:

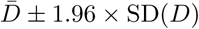

#### 2.8.3 Group comparisons and regression analyses

We explored the effects of imaging modality, time interval between two consecutive scans, stroke lesion size, demographic factors that have been reported to relate with WMH volume and/or brain size (i.e., age and sex), and WMH volume in the first scan, on segmentation performance.

In the MSS3 dataset, baseline imaging acquired at acute presentation (designated V0) consisted of either a diagnostic CT scan or a clinical MRI. All subjects subsequently underwent an MRI research scan at the first follow-up visit (V1), which occurred up to three months post-onset. Of the 229 participants in MSS3, at V0, 46 received a CT scan and 182 received a clinical MRI scan (Figure 2); all participants underwent MRI at V1. Manually-edited reference binary masks for WMH and stroke lesions were generated from MRI data at both visits as previously described, in the T2-weighted V1 image space for each subject. WMH segmentations obtained from V0 (either CT or MRI) scans were transformed into the corresponding V1 space to enable within-subject comparisons.

Two paired groups were established: CT–MRI (CT at V0 and MRI at V1; *n* = 46) and MRI–MRI (MRI at V0 and MRI at V1; *n* = 181). Within the MRI–MRI cohort, two scans were unpaired (one at V0 and one at V1), one pair did not co-register well, and eight pairs were excluded because the stroke lesion manual annotations did not match with the neuroradiological reports, resulting in *n* = 172 MRI–MRI pairs for analysis.

To assess the possible impact of imaging modality on WMH change estimates, voxel-wise categories of change were identified based on the difference between V1 and V0 WMH masks (V1-V0): remain (WMH at both time points), growth (WMH at V1 only), and regression (WMH at V0 only). Distributions of the corresponding volumes, specifically WMH_stable vol_, WMH_grow vol_, and WMH_shrink vol_, were compared between the two *independent* groups (CT–MRI vs. MRI–MRI) using the Wilcoxon rank-sum test (equivalent to the Mann–Whitney U test). The null hypothesis stated that the distributions of each component of WMH changes were the same for both groups.

The same non-parametric approach was used to compare the two groups with respect to the time between V0 and V1 scans, age, sex, baseline (V0) WMH volume, index stroke lesion volume, old stroke lesion volume, and incident infarct lesion volume observed at V1.

To identify factors associated with changes in WMH from V0 to V1 within each group, we fitted two general linear models (one per group). The change in WMH served as the dependent variable, with baseline WMH volume as the main predictor, adjusting for age, sex, inter-scan interval, index stroke lesion volume, and old stroke lesion volume.

We further evaluated the impact of the type and volume of stroke lesions (old and new) in the accuracy of the segmentations (DSC metric) also through two general linear models with DSC as outcome variable and as predictors: 1) old stroke lesion volume, type of stroke and reference WMH volume, and 2) new stroke lesion volume, type of stroke and reference WMH volume.

#### 2.8.4 Correlation with clinical visual scores and perivascular space burden

We used **Spearman’s Correlation Coefficient** (*ρ*; two-sided test) to evaluate the association between the volume of WMH (in the CT scans at V0) estimated by the best-performing model and the visual Fazekas scores (periventricular and deep) [12], estimated in the V1 MRI scan. As per above, the subsample for this evaluation was the CT–MRI group (n=48).

Enlarged perivascular spaces (PVS) are a neuroimaging marker of SVD and can co-occur with WMH [32], and are assessed in three clinically-relevant regions, namely basal ganglia (BG), centrum semiovale (CSO) and midbrain. High-density PVS burden may appear as hypodense foci on CT and could be misclassified as WMH, representing a potential source of false positives in CT-based WMH segmentation. We therefore used Spearman correlation to test the associations between PVS burden and regional WMH false positive metrics in the BG and CSO. BG and CSO ROI masks were constructed by combining left and right hemisphere masks (BG = left ∪ right; CSO = left ∪ right) derived from parcelling the brain using FreeSurfer (https://surfer.nmr.mgh.harvard.edu/).

Regional PVS density was computed as a percentage of ROI volume, 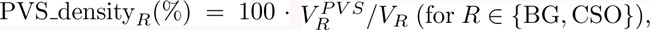 where 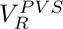 is the PVS volume within *R* and *V_R_* is the ROI volume [33]. Regional false positives were computed within each PVS ROI using the metrics defined in Section 2.8.1 (regional false positive burden and regional false positive concentration). Correlations were evaluated separately for BG and CSO; we report *ρ* and the corresponding p-value.

## 3 Results

### 3.1 Data Cleaning

After data cleaning and exclusions, 282 paired axial FLAIR-CT scans from three different datasets were included in this study: 91 from 80 MSS3 patients, 154 from 82 IST-3 patients, and 37 from the CIM dataset (see Figure 2).

### 3.2 MRI sequence identification

We manually annotated 162 MRI DICOM series (i.e., images) to validate the MRI sequence identification process. From them, 114 (70.4%) were correctly identified, 19 diffusion weighted images (11.7%) that had T2 contrast were identified as T2-weighted, therefore not considered entirely correct as these were not T2-weighted structural images. In 4 cases (2.5%) the dual-echo T2-weighted/proton density series were identified as proton density, which is not entirely correct either although can be acceptable. In 25 cases (15.4%) the structural sequences T1-weighted, T2-weighted or FLAIR were not identified, therefore labelled as “unknown”. However, whenever the FLAIR and the T1-weighted were identified, they were correctly labelled.

### 3.3 Preserving Anatomical Fidelity: Refining Registration and Architectural Design

We performed a series of experiments to refine both the preprocessing pipeline and the model architecture for CT-based WMH segmentation. We established a baseline model by training a standard 3D nnU-Net on native CT images (Experiment 1). For this baseline model, WMH ground truth masks originally delineated on MRI/FLAIR were transferred to the CT space through a single-step rigid-body registration using FSL FLIRT [26]. This configuration attained an average DSC of 0.562, average sensitivity of 0.526, a MAE in WMH volume estimation of 3.696 mL, and an average precision of 0.634 (Table IV).

**Table IV:** Segmentation performance metrics (mean ± standard deviation) averaged over 5-fold cross-validation across various experiments. DSC = Dice similarity coefficient; MAE = mean absolute error.

| Model | Experiment Description | DSC | Sensitivity | MAE (mL) | Precision |
| --- | --- | --- | --- | --- | --- |
| 1 | Baseline model with native CT images | $0.562 \pm 0.167$ | $0.526 \pm 0.168$ | $3.696 \pm 4.199$ | $0.634 \pm 0.177$ |
| 2 | CT images registered to T1-weighted MRI template | $0.545 \pm 0.172$ | $0.513 \pm 0.171$ | $3.993 \pm 4.825$ | $0.622 \pm 0.193$ |
| 3 | CT images registered to CT template | $0.548 \pm 0.168$ | $0.513 \pm 0.168$ | $5.577 \pm 6.832$ | $0.626 \pm 0.191$ |
| 4 | nnU-Net with Residual Blocks using FSL (rigid registration) | $0.563 \pm 0.167$ | $0.523 \pm 0.168$ | $3.566 \pm 4.077$ | $0.640 \pm 0.177$ |
| 5 | nnU-Net with Residual Blocks using NiftyReg (rigid + affine registration) | $0.567 \pm 0.169$ | $0.527 \pm 0.169$ | $3.497 \pm 3.836$ | $0.646 \pm 0.178$ |

In Experiments 2 and 3 we investigated the effect of registering CT images to standard anatomical templates, as previously suggested by [8] and [9]. When CT scans were aligned to a T1-weighted MRI template (Experiment 2) or a CT template (Experiment 3), segmentation performance was slightly poorer than the baseline model. The MAEs were higher (that is, 3.99 mL for Experiment 2 and 5.58 mL for Experiment 3), and the DSCs were lower (that is, 0.545 and 0.548, respectively). These findings indicate that key anatomical features required for WMH delineation are disrupted by template-based registration. Interpolation and resampling smooth subtle lesions, or introduce artefacts, ultimately degrading segmentation quality. Therefore, preserving native CT spatial characteristics is important to retain robust WMH segmentation.

In Experiment 4, we explored architectural refinements aiming to improve the model’s feature extraction and stability. The network architecture for Experiment 4, is built upon the baseline model (Experiment 1) trained on native CT images, by adding residual blocks to the nnU-Net. Aligned with current state-of-the-art practices [34], this modification slightly improved the WMH volume estimation (MAE reduced from 3.696 mL to 3.566 mL), suggesting that residual connections improved feature learning and gradient flow.

In Experiment 5, we combined the architectural improvement of Experiment 4 with a refined registration to further improve the alignment of WMH masks between FLAIR and CT images. In this experiment, instead of relying on rigid registration, we applied rigid and affine transformations using NiftyReg. This approach gave us a slightly better DSC of 0.567 and a smaller MAE (3.497 mL).

In addition to calculating the DSC and MAE by comparing the results obtained with the ground truth in the native CT space, we transferred the results of the segmentation to the FLAIR native space and calculated the MAE in this space to evaluate the effect of the added space transformation in the segmentation task. In this evaluation, the MAE from Experiment 4 was 3.831, and for Experiment 5 it was 3.705. Although Experiment 5 yielded better numeric results, the MAE in both was higher than when all masks were in the CT space (IV). These results demonstrate that a nuanced geometric refinement is required for the desired mask alignment.

In summary, template-based registration was found to be detrimental to maintaining important native CT features necessary for accurate segmentation. Refining mask alignment with affine transformations improved overall segmentation accuracy and enabled more reliable CT-based WMH assessment.

### 3.4 Fine-Tuning with Pseudo-Labelled Data Improves WMH Segmentation and Volume Estimation

We observed that the segmentation performance of the model in Experiment 5 was particularly poor in cases with mild WMH burden (WMH volume ≤ 10 mL), as illustrated in Figure 6, indicating a need for further refinement to improve the model’s performance for subtle WMH regions.

**Figure 6:**
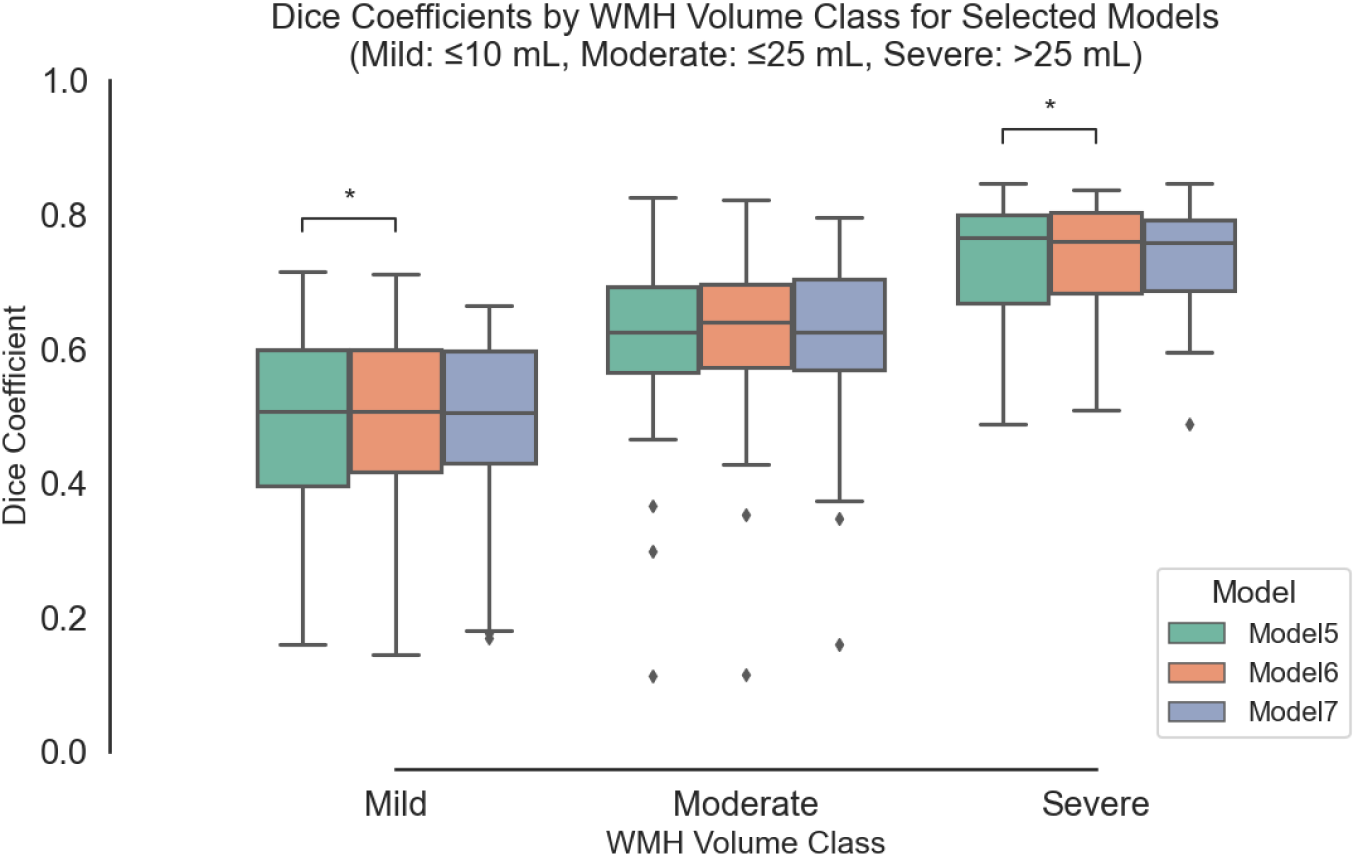
The distribution of Dice coefficients across three WMH volume classes for the models in experiments 5, 6, and 7. The comparison highlights the segmentation performance of each model within mild, moderate, and severe WMH burden categories. Model 7 demonstrates slightly better performance in mild and severe WMH volume classes.

A possible reason for the suboptimal performance in segmenting mild WMH is the low soft tissue contrast in CT, which makes it difficult for the model to separate these faint lesions from adjacent normal white matter tissue or image noise. This, in addition to their variation in shape, and location, makes it difficult for the model to learn consistent spatial features accurately representing subtle and small lesions. We explored whether adding more training examples with mild WMH could enable the model to learn more specific features of these lesions.

Hence, we fine-tuned the model used in Experiment 5 with additional pseudo-labelled data from the CIM dataset, which included healthy individuals with mild age-related WMH (Experiment 6). Although the overall Dice scores and MAE showed slight improvement, the improvement in Dice scores and MAE specifically for cases with mild WMH burden was almost negligible (see Table V, Figures 6 and 7). This reflects the inherent difficulty in accurately segmenting mild WMH lesions. The challenge likely arises not only from data scarcity but also from their low tissue contrast, which fades in CT making these small lesions difficult to differentiate from adjacent normal tissue or noise.

**Figure 7:**
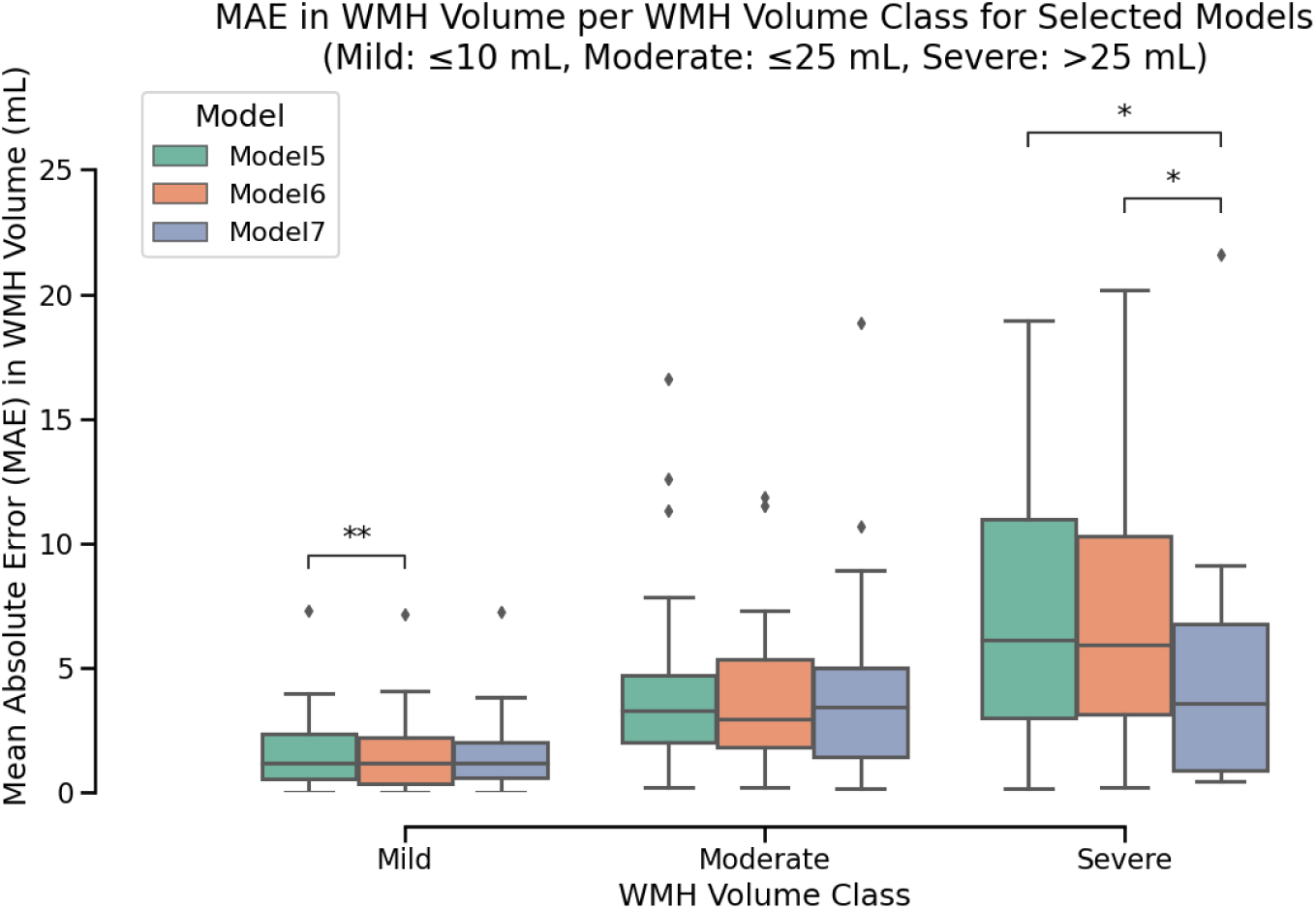
The distribution of Mean Absolute Error (MAE) values for WMH volume estimation across three WMH volume classes (Mild: *≤*10 mL, Moderate: *≤*25 mL, Severe: *>*25 mL) for Models 5, 6, and 7. The fine-tuned model (Model 7) demonstrates significantly improved performance in the severe WMH volume class (*p <* 0.05) compared to Model 5.

**Table V:** Segmentation performance metrics (mean ± standard deviation) averaged over 5-fold cross-validation before and after fine-tuning. DSC = Dice similarity coefficient; MAE = mean absolute error.

| Model | Experiment Description | DSC | Sensitivity | MAE (mL) | Precision |
| --- | --- | --- | --- | --- | --- |
| 5 | nnU-Net with Residual Blocks using NiftyReg (rigid + affine registration) | $0.567 \pm 0.169$ | $0.527 \pm 0.169$ | $3.497 \pm 3.836$ | $0.646 \pm 0.178$ |
| 6 | Fine-tuning Model 5 with CIM dataset | $0.575 \pm 0.165$ | $0.534 \pm 0.168$ | $3.229 \pm 3.755$ | $0.651 \pm 0.169$ |
| 7 | Fine-tuning Model 5 with both CIM and IST-3 datasets | $0.569 \pm 0.165$ | $0.546 \pm 0.166$ | $2.928 \pm 3.519$ | $0.620 \pm 0.175$ |

We further fine-tuned with additional pseudo-labelled data from the IST-3 dataset (Experiment 7) to expand the diversity of WMH presentations exposed to the model. The overall DSC slightly improved compared to the model in Experiment 5 (see Table V). Additionally, a slight but consistent increase in sensitivity (0.546), (see Table V) was observed with fine-tuning. This suggests that pseudo-labelled data helps the model identify a slightly higher proportion of WMH regions. The MAE for volume estimation was also reduced to 2.93 mL (Table V). This implies that the last enhanced model was more generalisable and more proficient at detecting WMH lesions with various degrees of severity. We also observed significant improvement in Dice and MAE in severe WMH cases (Figures 6 and 7), which demonstrates the value of using multiple datasets, even pseudo-labelled, to improve model robustness.

Figures 8 and 9 show the effect of pseudo-labelled data (model from Experiment 7) versus training only with manually labelled data (model from Experiment 5). The difference in Dice coefficients (Model 7 - Model 5) across mild, moderate, and severe WMH classes is shown in Figures 8. Positive values indicate better overlap accuracy in Model 7, while negative values favour Model 5. The median DSC difference for mild and severe cases are near zero, showing slight improvement, but Model 7 demonstrates slightly better consistency for moderate cases. The difference in MAE for volume estimation is shown in Figure 9. Here, negative values indicate lower MAE for Model 7, i.e. better volume accuracy. Performance is similar for mild cases, but Model 7 has slightly better accuracy for moderate cases and a considerable improvement for severe cases, likely attributable to the pseudo-labelled data adding more diverse WMH examples to the training data. These results emphasise the challenges of segmenting mild lesions and the benefits of fine-tuning for severe WMH.

**Figure 8:**
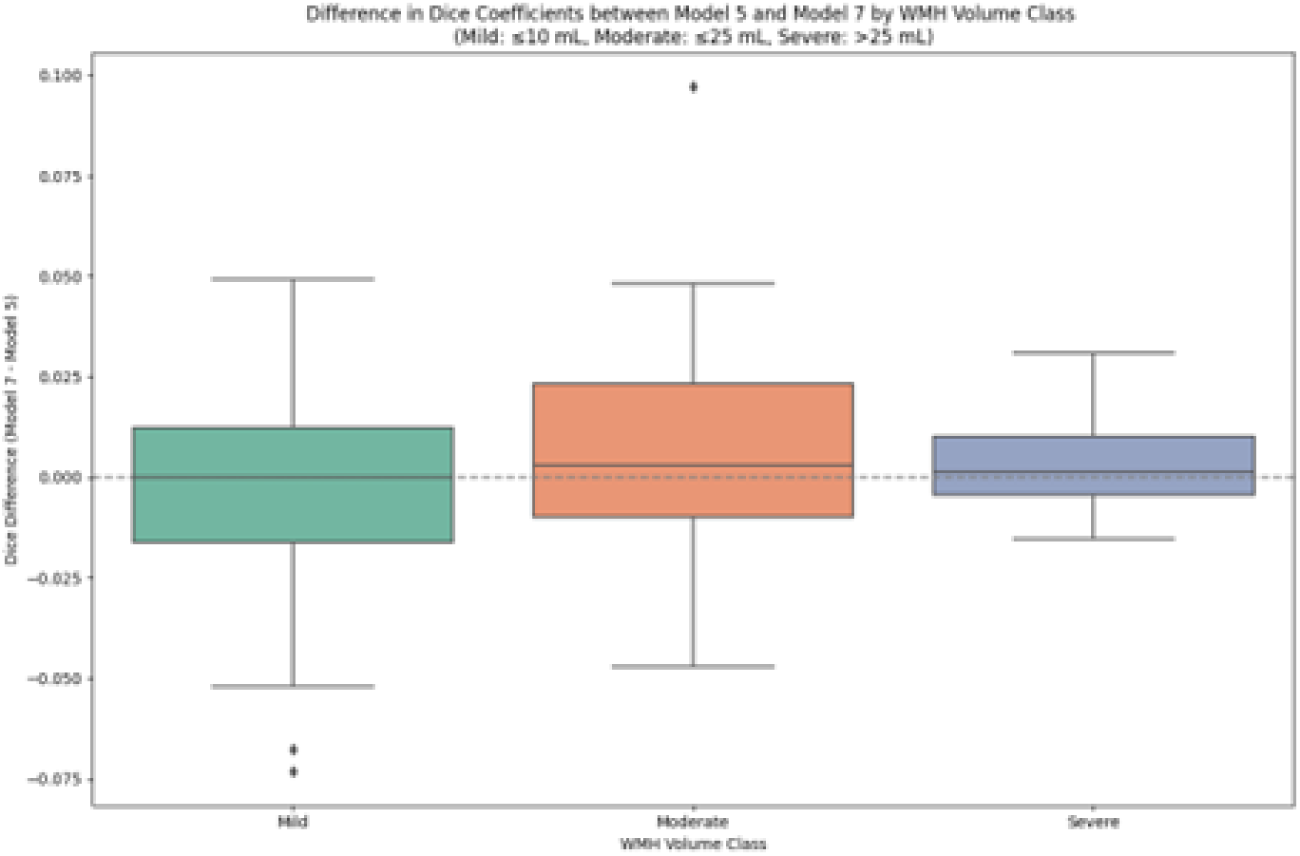
Difference in Dice Coefficients between Model 5 and Model 7 by WMH Volume Class (Mild, Moderate, Severe); (Model 7 - Model 5)

**Figure 9:**
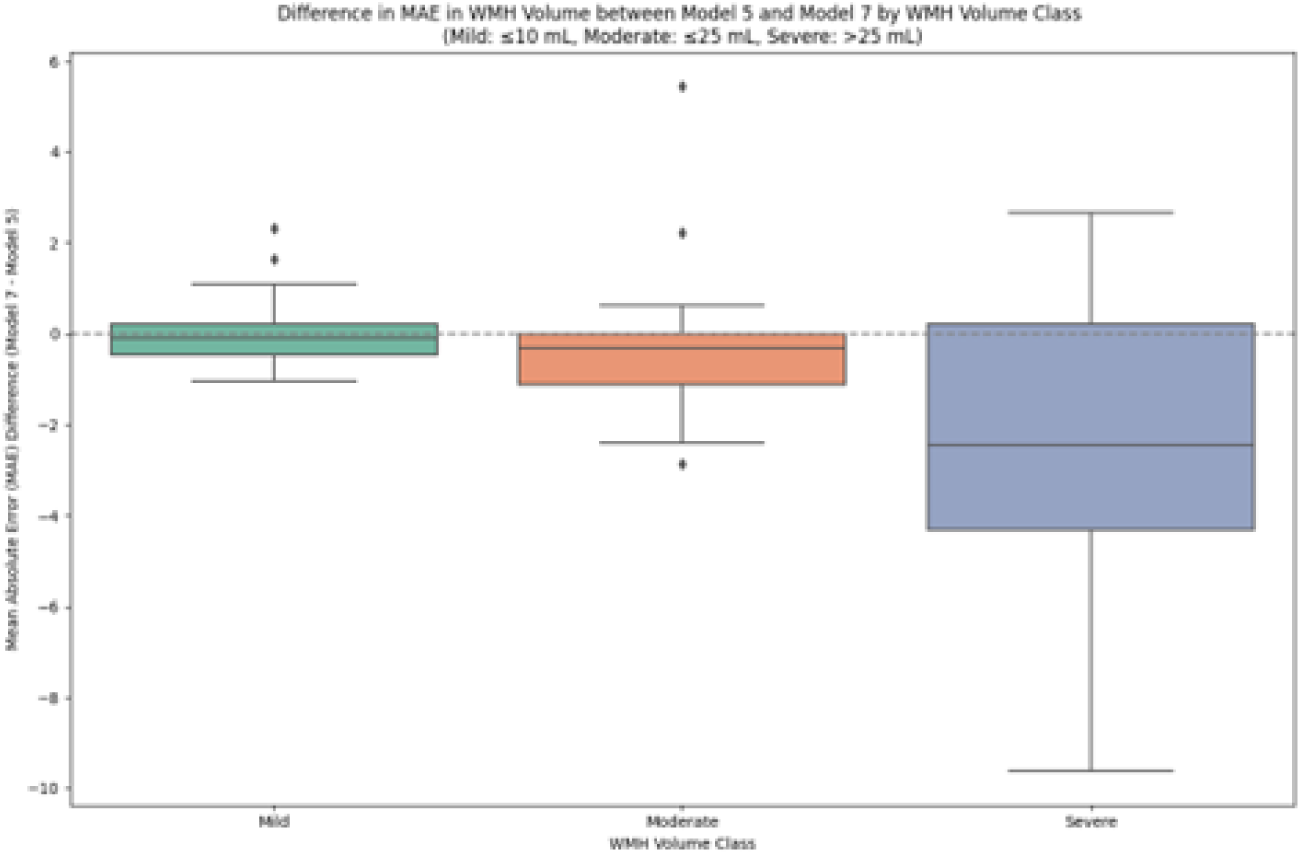
Difference in Mean Absolute Error (MAE) in WMH Volume between Model 5 and Model 7 by WMH Volume Class (Mild, Moderate, Severe); (Model 7 - Model 5)

Figure 12 provides qualitative examples of the effect of pseudo-label fine-tuning (Model 7) relative to the baseline model (Model 5) across three cases spanning WMH burden: one mild case (GT WMH volume ≤10 mL) and two moderate cases (GT WMH volume *>* 10–≤25 mL). As illustrated, the finetuned model reduces fine-lining periventricular and scattered deep white-matter false positives while largely preserving major confluent WMH regions. These examples are consistent with the quantitative trend towards improved precision and reduced false-positive burden after fine-tuning.

The performance of the fine-tuned nnUNet model (Model 7) was further validated by comparing the WMH volumes derived from CT segmentations with ground truth (GT) FLAIR segmentations. Figure 10 (left) shows the correlation between the nnUNet-predicted WMH volumes derived from CT-based segmentations and ground-truth (GT) MRI WMH volumes registered to CT space (*r* = 0.9853, *p <* 0.00001).

**Figure 10:**
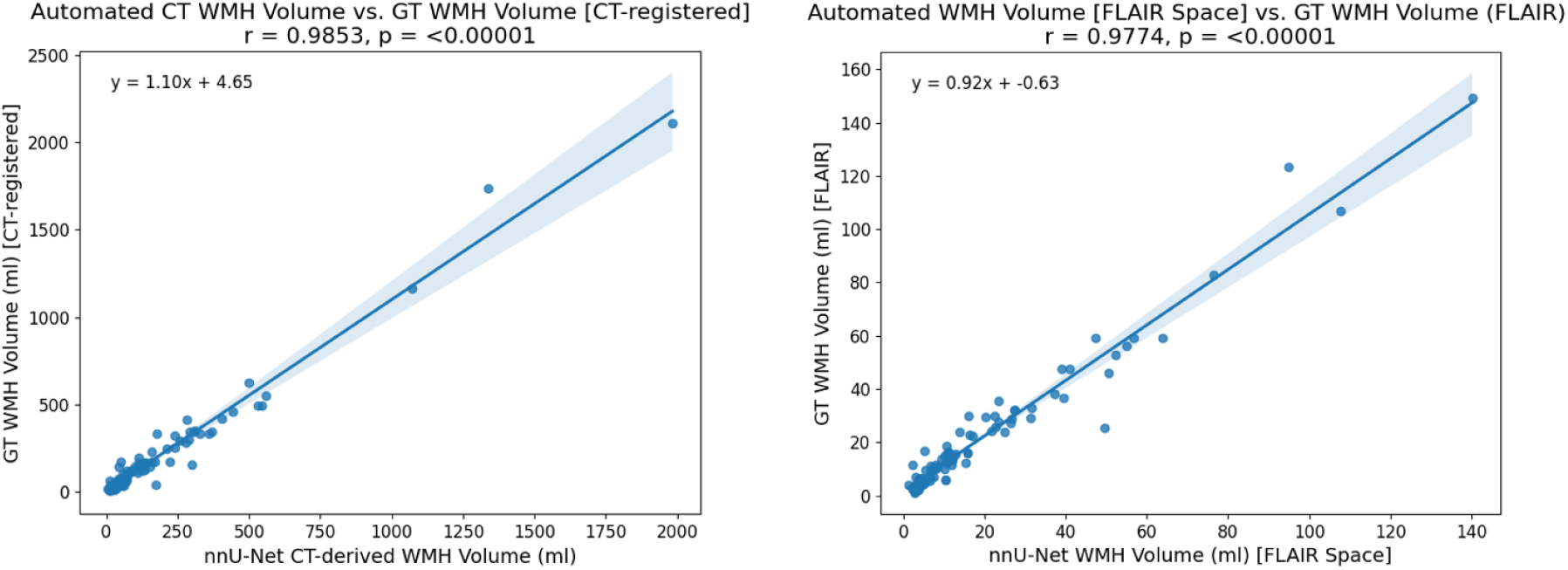
(Left) Correlation between nnUNet-predicted WMH volumes derived from CT-based segmentations and ground-truth (GT) MRI WMH volumes registered to CT space (*r* = 0.9853, *p <* 0.00001). (Right) Correlation between nnUNet-predicted WMH volumes and GT MRI WMH volumes, both evaluated in native FLAIR space (*r* = 0.9774, *p <* 0.00001). Both analyses demonstrate high linear agreement, indicating robust model performance across spatial domains.

The correlation of nnUNet-predicted WMH volumes and GT MRI WMH volumes, both evaluated in native FLAIR space is shown in Figure 10 (right). A strong correlation (*r* = 0.9774, *p <* 0.00001).

The Bland-Altman plot (Figure 11) illustrates the differences between the FLAIR reference volumes and nnUNet-predicted CT WMH volumes (in FLAIR space). The mean difference is slightly above zero, which suggests an underestimation of WMH volumes (in average) by the nnUNet model. The differences close and outside the confidence intervals typically correspond to complex anatomical disruptions such as large stroke lesions, irregular shapes, or low tissue contrast. This highlights the model’s limitations in segmenting WMH in these difficult cases.

**Figure 11:**
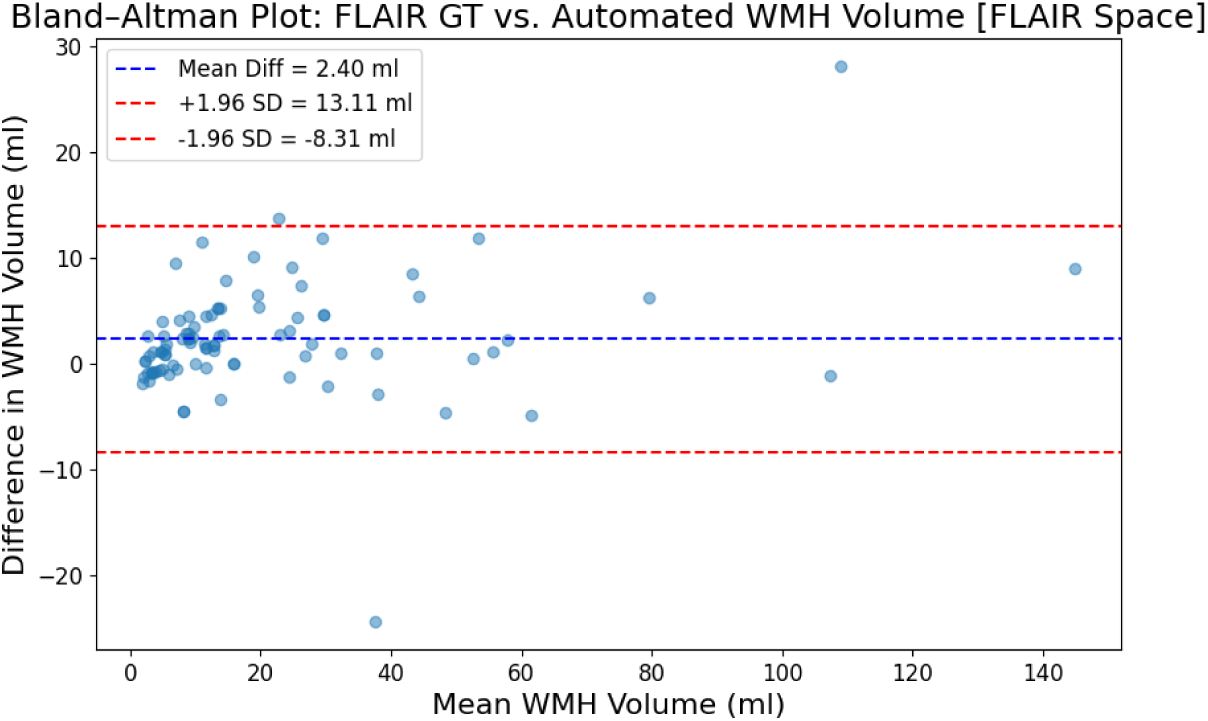
Bland–Altman plot illustrating the agreement between automated nnUNet–predicted WMH volumes and ground-truth (GT) MRI WMH volumes evaluated in native FLAIR space. The mean difference (bias) was 2.40 mL (blue dashed line), with 95% limits of agreement ranging from -8.31 mL to 13.11 mL (red dashed lines).

**Figure 12:**
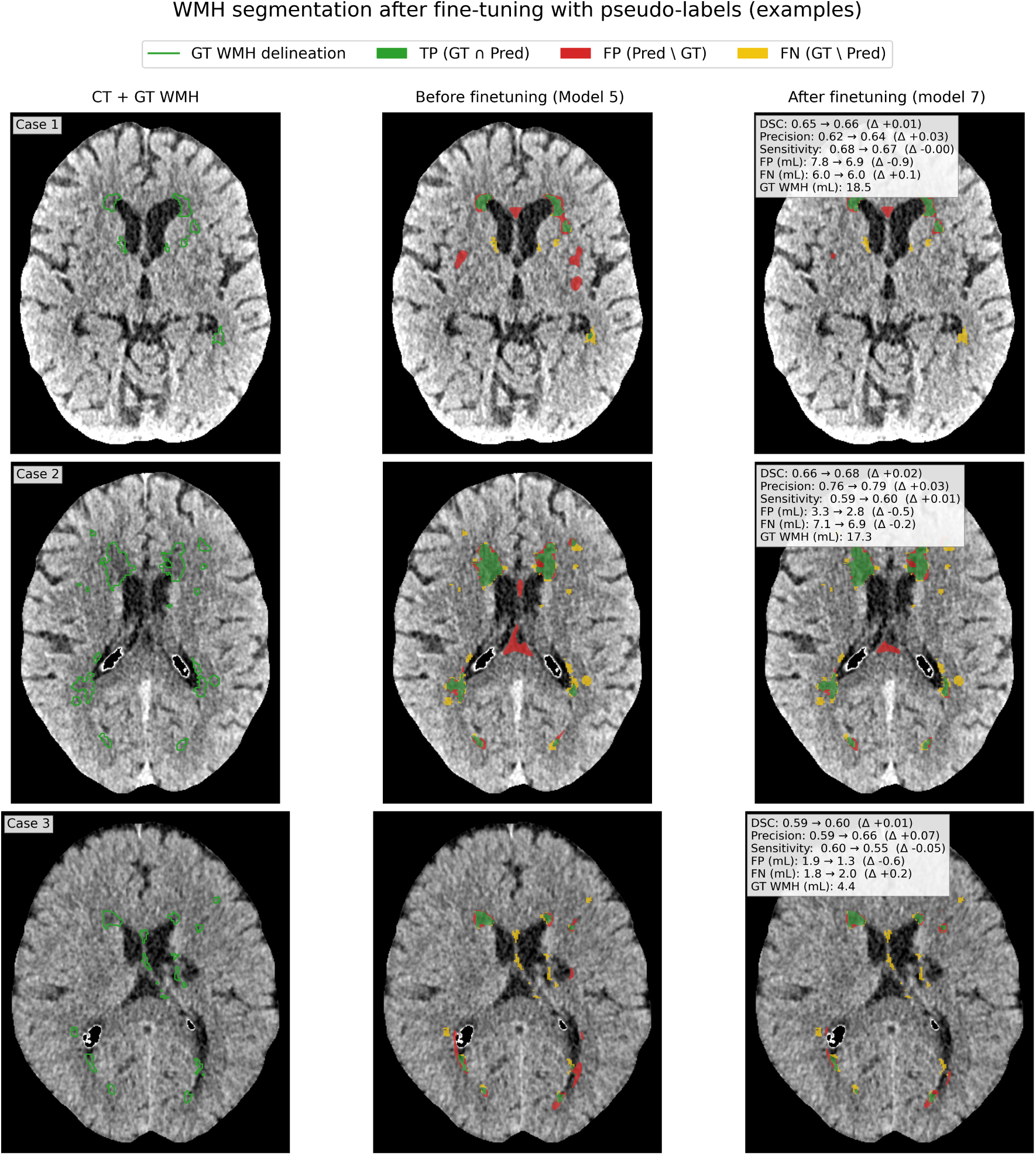
Qualitative examples of WMH segmentation changes after finetuning with pseudo-labels. Three example cases are shown in CT space. Left: CT with fluid-attenuated inversion recovery (FLAIR)-derived WMH *ground truth* (GT) delineation (green) Middle: Model 5 error map. Right: fine-tuned model (Model 7) error map. In the error maps, *true positives* (TP; GT∩Pred) are shown in green, *false positives* (FP; Pred\GT) in red, and *false negatives* (FN; GT\Pred) in yellow, where Pred denotes the model *prediction*. Insets report Dice similarity coefficient (DSC), precision, and sensitivity (recall), together with false-positive (FP) and false-negative (FN) volumes in millilitres (mL). In the insets, values are reported as baseline → fine-tuned (Model 5 → Model 7), and Δ denotes the change from baseline to fine-tuned (Model 7−Model 5).

These results generally show that the fine-tuned model is reliable in estimating WMH volumes with good correlation and concordance between CT and FLAIR-based segmentations.

### 3.5 Scanning Modality, Not Time Intervals, Significantly Affects WMH Volume Estimation

CT-based V0 assessments systematically differed from the MRI-based ones, likely due to intrinsic differences in contrast, spatial resolution, and segmentation accuracy. Comparisons between the CT-MRI (n=46) and MRI-MRI (n=172) groups revealed substantial differences in WMH changes. The Mann-Whitney U test revealed statistically significant disparities across five examined variables (WMH_shrinkvol_, WMH_grow vol_, WMH volume difference (V1-V0), age of the participants in each patient group, and index stroke lesion volume), with p-values *<* 0.0001 for WMH_grow vol_, WMH volume difference (V1-V0), and index stroke lesion volume, p=0.0020 for age and p=0019 for WMH_shrink vol_. However, the distribution of time differences between V0 and V1 scans was not different in the CT-MRI and MRI-MRI patients groups (p=0.1119) (Figure 13). The sex (p=0.9472), old stroke lesion volume (p=0.2050), and volume of recent infarct lesions (p=0.8287) did not differ between the two patients groups either.

**Figure 13:**
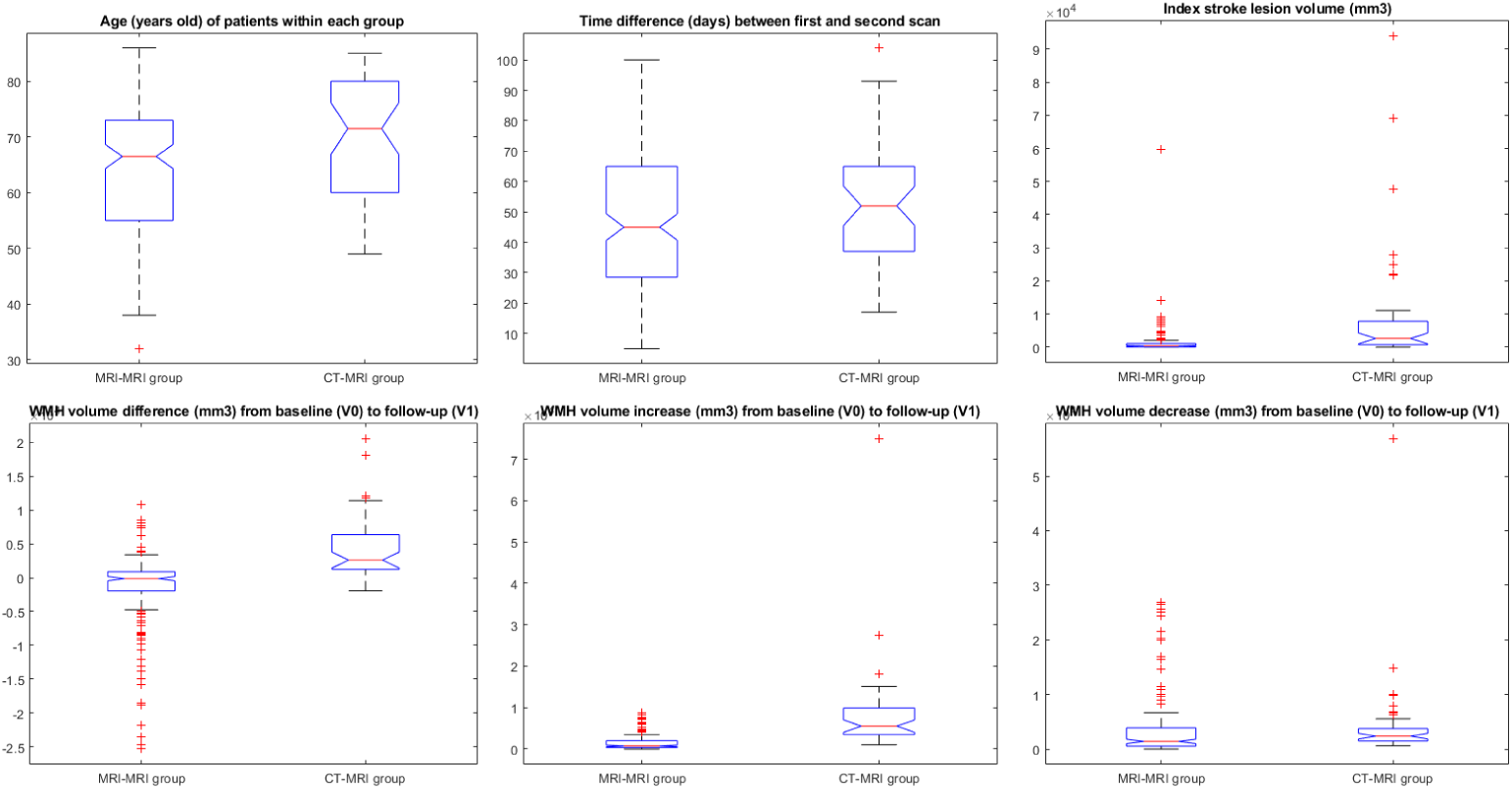
Box plots of age, time difference between first and second scan, index stroke lesion volume, WMH volume difference (V1-V0), WMH_grow vol_, and WMH_shrink vol_ for the CT-MRI and MRI-MRI patients groups.

### 3.6 Previous stroke lesion volume and time between scans influence the WMH volume difference only in MRI measurements

As Table VI shows, the initial volume of WMH (at V0), the volume of previous (that is, old) stroke lesions, and the time between scans, were influential in the WMH volume difference between the V0 and 3-month follow-up (i.e., V1) WMH volumes in the MRI-MRI patient group, while in the CT-MRI group only the WMH volume at V0 influenced the change in WMH volume between V0 and V1.

**Table VI:** Results from general linear models evaluating drivers of WMH change (outcome variable) between the two time points for each group. *Groups:* CT-MRI = baseline CT and follow-up MRI; MRI-MRI = baseline MRI and follow-up MRI. WMH: white matter hyperintensities (MRI) / hypoattenuations (CT); SL: stroke lesion; Vol: volume. Volumes are in *mm*^3^ and age is in years.

| Group | Predictor | B-value | Std. error | p-value | Significant |
| --- | --- | --- | --- | --- | --- |
| CT-MRI | WMH Vol (V0) | 0.1800 | 0.0311 | $p < 0.0001$ | **Yes** |
| CT-MRI | Age (V0) | -107.90 | 60.4870 | 0.0822 | No |
| CT-MRI | Sex | 1103.4 | 1254.5 | 0.3845 | No |
| CT-MRI | Time (days) | -36.698 | 27.74 | 0.1935 | No |
| CT-MRI | index SL Vol | 0.0284 | 0.0307 | 0.3603 | No |
| CT-MRI | old SL Vol | 0.0878 | 0.0953 | 0.3620 | No |
| MRI-MRI | WMH Vol (V0) | -0.1351 | 0.0238 | $p < 0.0001$ | **Yes** |
| MRI-MRI | Age (V0) | 31.043 | 38.073 | 0.4160 | No |
| MRI-MRI | Sex | -951.08 | 847.39 | 0.2633 | No |
| MRI-MRI | Time (days) | -42.475 | 16.603 | 0.0114 | **Yes** |
| MRI-MRI | index SL Vol | -0.0464 | 0.0805 | 0.5651 | No |
| MRI-MRI | old SL Vol | 0.2365 | 0.1197 | 0.0497 | **Yes** |

### 3.7 Impact of Stroke Lesions on WMH Segmentation

The presence of historical (old) and current (index) stroke lesions, each with different sizes and complexities, made it difficult to automate WMH delineation. Mean old stroke lesion volumes in the MSS3 dataset were 1,044.68 mm^3^ (SD = 4,459.70 mm^3^), and mean index stroke lesion volumes were larger: 5,254.91 mm^3^ (SD = 13,665.16 mm^3^). The mean WMH volume was 17,259 mm^3^ (SD = 20,138 mm^3^) and there was a wide range of lesion volumes within the cohort.

Moderate decrease in Dice scores was associated with larger old stroke lesion volumes (standardised beta = -0.2319; p = 0.015), indicating that historical structural changes (presumably scarring and tissue remodelling) mildly complicated WMH delineation. However, index stroke lesion volumes had a more pronounced negative effect (standardised beta = -0.3075, *p <* 0.001), suggesting that the presence of recent ischaemic injury (oedema, heterogeneous tissue composition, altered local intensity) was more difficult for the model to deal with (Supplementary Figure 3). Conversely, the volume of WMH lesions positively affected segmentation accuracy (standardised beta = 0.6791; *p <* 0.001), indicating that higher WMH volumes result in more explicit lesion boundaries and, therefore, more accurate delineations.

Specific stroke types (e.g., cortical, lacunar, subcortical) generally had a smaller influence. Other stroke types did not significantly affect Dice scores, but old lacunar stroke lesions negatively affected segmentation accuracy (standardised beta = -0.9948; p = 0.049). The current (index) stroke subtype also did not meaningfully affect segmentation performance, suggesting that lesion volume and complexity, rather than morphological stroke subtype, are more important determinants of accuracy.

### 3.8 Correlation with visual ratings and perivascular space burden

WMH burden estimated by the best-performing model showed strong correlations with visual Fazekas grades. Predicted CT WMH burden correlated with periventricular Fazekas grade (*ρ* = 0.825, *p* = 5.66 × 10*^−^*^13^) and deep Fazekas grade (*ρ* = 0.818, *p* = 1.28 × 10*^−^*^12^). Ground-truth (i.e., FLAIR-derived) WMH burden showed similarly strong correlations (periventricular: *ρ* = 0.818, *p* = 1.21 × 10*^−^*^12^; deep: *ρ* = 0.769, *p* = 1.76 × 10*^−^*^10^); see Table VII.

**Table VII:** Spearman correlations between Fazekas visual ratings and WMH burden (*n* = 48). *Predictors:* PV Fazekas = periventricular Fazekas grade; DWM Fazekas = deep white-matter Fazekas grade. *Outcomes:* Predicted WMH volume = modelderived WMH volume; GT WMH volume = FLAIR-derived WMH volume.

| Predictor | Outcome | $\rho$ | $p$ |
| --- | --- | --- | --- |
| PV Fazekas | Predicted WMH volume | 0.8249 | $5.66 \times 10^{-13}$ |
| PV Fazekas | GT WMH volume | 0.8184 | $1.21 \times 10^{-12}$ |
| DWM Fazekas | Predicted WMH volume | 0.8179 | $1.28 \times 10^{-12}$ |
| DWM Fazekas | GT WMH volume | 0.7688 | $1.76 \times 10^{-10}$ |

PVS burden was correlated with WMH volume of false positives in BG and CSO. Regional WMH volume of false positive (in number of voxels) increased with regional PVS density in BG (*ρ* = 0.556, *p* = 4.08 × 10*^−^*^5^) and CSO (*ρ* = 0.416, *p* = 3.24 × 10*^−^*^3^). When WMH volumes of false positives were normalised by ROI volume (false positives per mL; voxels/mL), the correlation remained significant (BG: *ρ* = 0.529, *p* = 1.12 × 10*^−^*^4^; CSO: *ρ* = 0.424, *p* = 2.69 × 10*^−^*^3^; Figure 14). In addition, the proportion of false positives in the ROIs with respect to the total of false positives, also correlated highly with the PVS density measurements (BG FP concentration: *ρ* = 0.388, *p* = 6.39 × 10*^−^*^3^; CSO FP concentration: *ρ* = 0.348, *p* = 1.52 × 10*^−^*^2^), and overall *ρ* = 0.366, *p* = 1.04 × 10*^−^*^2^). Representative overlays illustrating FP localisation within the PVS ROIs are shown in Fig. 15. All correlations between PVS density and the WMH false-positive metrics are summarised in Table VIII.

**Figure 14:**
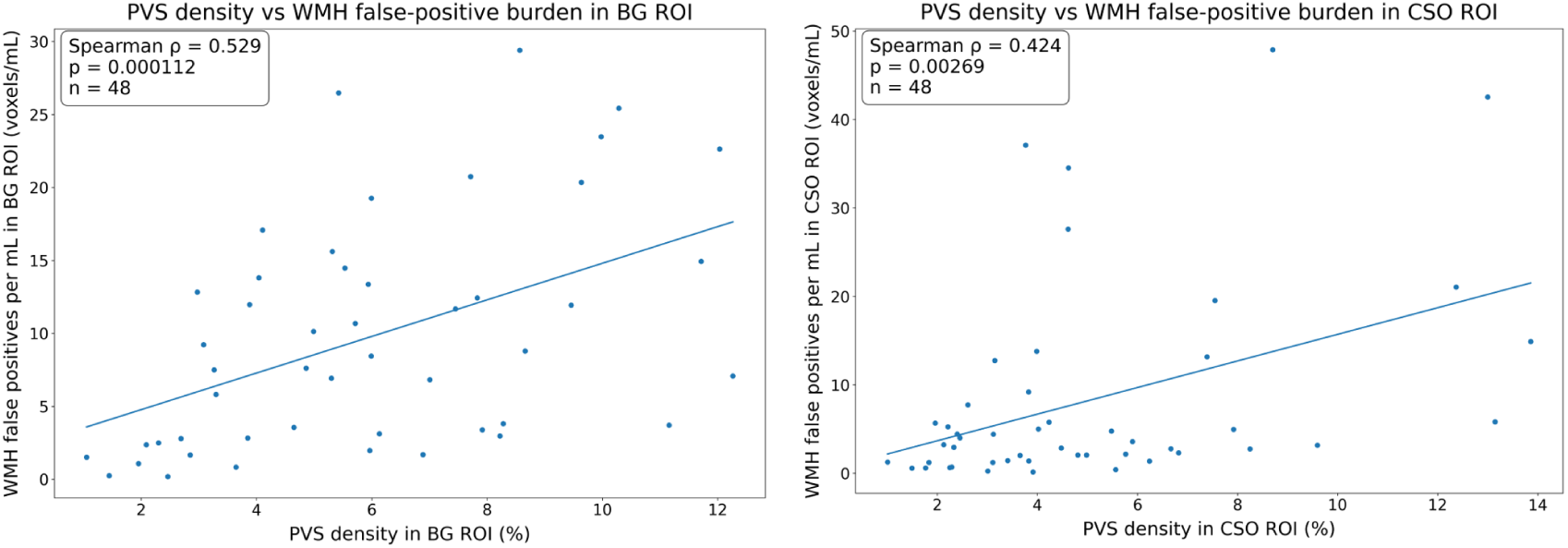
Association between PVS density (%) and WMH false-positive burden normalised by ROI volume (voxels/mL) in basal ganglia (BG; left) and centrum semiovale (CSO; right). Spearman’s *ρ*, p-value, and sample size (*n*) are reported in each panel.

**Figure 15:**
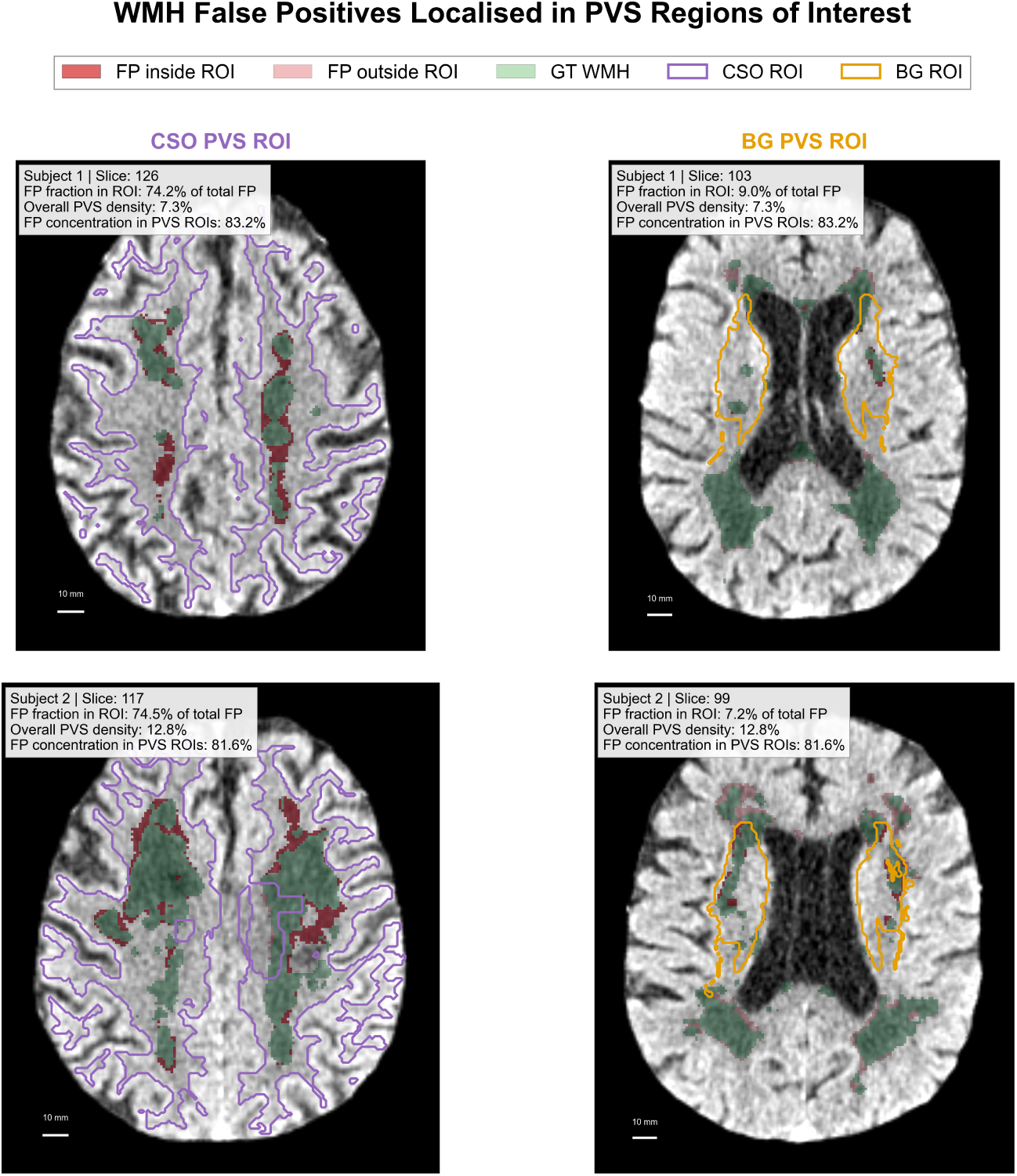
Example visualisation of WMH false positives (FP) within perivascular space (PVS) regions of interest (ROIs) in the centrum semiovale (CSO) and basal ganglia (BG) for two representative subjects. FP voxels inside the ROI are shown in dark red and FP voxels outside the ROI are shown in light red; FLAIR-derived WMH ground truth is overlaid in green. CSO and BG ROIs are outlined in purple and orange, respectively. Text boxes report the ROI FP fraction (percentage of total FP), overall PVS density, and overall FP concentration within the combined PVS ROIs.

**Table VIII:**
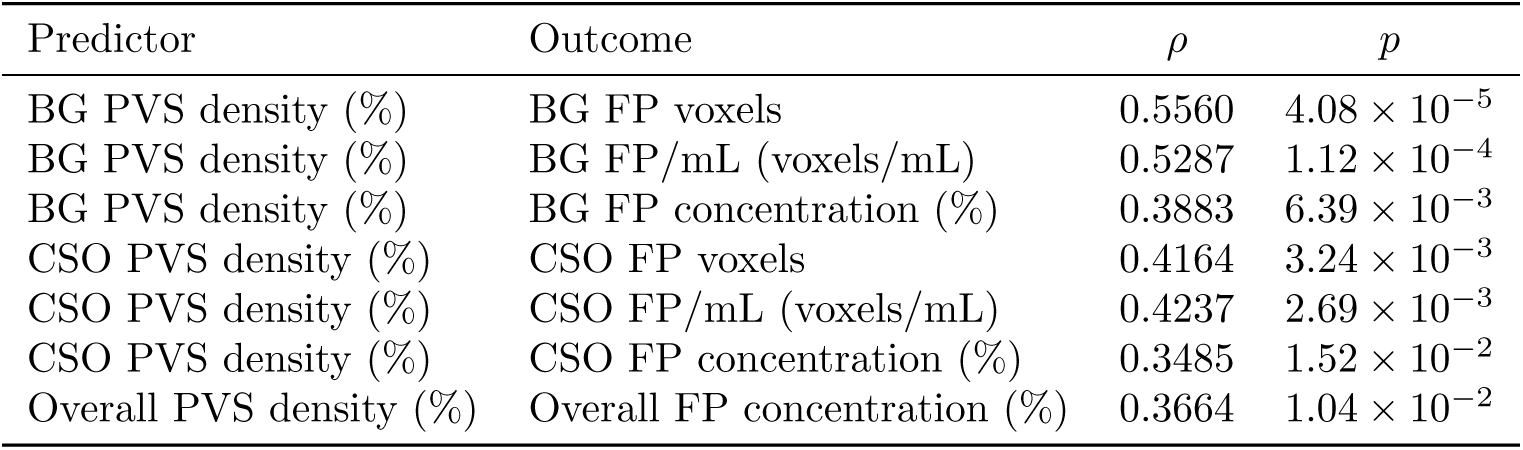
Spearman correlations between PVS density and regional WMH falsepositive (FP) metrics (*n* = 48). PVS density is reported as % of ROI volume (BG = basal ganglia; CSO = centrum semiovale). FP voxels are defined as Pred ∩ ¬GT. FP/mL denotes FP voxels normalised by ROI volume (voxels/mL). FP concentration denotes the proportion of total FP located within the specified ROI. Overall FP concentration denotes (FP*_BG_* + FP*_CSO_*)*/*FP*_total_*.

| Predictor | Outcome | $\rho$ | $p$ |
| --- | --- | --- | --- |
| BG PVS density (%) | BG FP voxels | 0.5560 | $4.08 \times 10^{-5}$ |
| BG PVS density (%) | BG FP/mL (voxels/mL) | 0.5287 | $1.12 \times 10^{-4}$ |
| BG PVS density (%) | BG FP concentration (%) | 0.3883 | $6.39 \times 10^{-3}$ |
| CSO PVS density (%) | CSO FP voxels | 0.4164 | $3.24 \times 10^{-3}$ |
| CSO PVS density (%) | CSO FP/mL (voxels/mL) | 0.4237 | $2.69 \times 10^{-3}$ |
| CSO PVS density (%) | CSO FP concentration (%) | 0.3485 | $1.52 \times 10^{-2}$ |
| Overall PVS density (%) | Overall FP concentration (%) | 0.3664 | $1.04 \times 10^{-2}$ |

## 4 Discussion

We present an end-to-end AI-powered framework for WMH segmentation in non-contrast brain CT scans that leverages the use of a state-of-the-art deep learning configuration and a combination of manually annotated and pseudo-labelled CT-MRI/FLAIR paired data. By integrating research-quality manual annotations from the MSS3 dataset with pseudo-labelled data derived from routine clinical data from the IST-3 and out-of-distribution data from the CIM cohort, our core approach addressed two longstanding challenges in CT-based WMH segmentation: (1) the relative underperformance of CT-based methods compared to the MRI-based counterparts, and (2) the limited explainability of the state-of-the-art models developed for the same task, which have hindered the increase of the performance of these models.

### 4.1 Data Preparation

Processing raw clinical image data is challenging. Information from DICOM tags is inconsistent, sometimes missing or incorrect, and image data is very heterogeneous in terms of quality, signal-to-noise ratio, and acquisition settings. By using a two-way complementary approach to identify the image data for consistently labelling and organising it in BIDS-NIfTI-1 format, we dealt with the heterogeneity of the information in the relevant DICOM tags. Still, visual inspection was needed in some cases to discern usability between images of the same type. If larger datasets would have been used, automatic ways to check the data preparation pipeline would have been necessary. For example, image quality could have been appraised using mriqc https://mriqc.readthedocs.io/en/stable/, and brain coverage could have been explored following the approach described by Jin et al. [35], available from https://github.com/bjin96/ct-processing.

Importantly, our analyses revealed that spatial normalisation to standard MRI or CT templates diminished segmentation performance. Registering CT scans to such templates introduced interpolation effects and disrupted subtle intensity cues crucial for identifying WMHs. This observation contrasts with previous CT-based attempts ([8], [9]), which employed registration to MRI or CT templates and reported relatively modest DSC of about 0.43 and correlation with Fazekas scores. Our findings suggest that preserving native CT spatial properties is important for maintaining tissue contrast and lesion visibility, which may help explain why template registration may have constrained performance in prior work.

### 4.2 Data Integration

The incorporation of pseudo-labelled training data improved both the model’s robustness and accuracy, positively impacting the volumetric estimation. Sensitivity increased from 0.527 to 0.546, and MAE in the estimation of the WMH volume decreased from 3.497 mL to 2.928 mL. The improvement was especially visible in cases with a high WMH burden, where exposure to a broader range of lesion appearances helped the network to delineate subtle and severe lesions equally well.

Previous efforts have consistently reported lower segmentation accuracy and limited generalisability when applying automated WMH segmentation methods to CT data, especially when compared to MRI-based approaches. For instance, Chen et al. [13] found a lower correlation (0.71) for CT compared to MRI (0.85) for the automated measurement of WMH volume. Similarly, Pitkänen et al. [8] achieved a DSC of 0.43, but their validation strategy was confined to a single dataset, raising concerns about overfitting and the model’s applicability in broader clinical contexts. Kaipainen et al. [9] reported good correlations with Fazekas scores; however, they did not provide spatial overlap measures or externally validate the segmentation model, limiting insights into the model’s true generalisability. Recent work by van Voorst et al. [15] showed promising results on internal data but had a significant drop in performance (DSC 0.23) on external validation.

In contrast, our approach achieved higher DSC scores (up to 0.575) and near-perfect CT-MRI volume correlations (*r* = 0.9805) despite incorporating a substantial proportion of mild WMH cases —around half of our validation samples — known to yield inherently lower DSC values due to subtle lesion appearances. Previous studies often focused on datasets with more balanced or moderately burdened WMH distributions, which can inflate performance metrics. Our ability to maintain high segmentation accuracy even under such challenging conditions suggests that integrating diverse datasets and leveraging pseudo-labelled training examples from MRI-based WMH models can yield a more generalised model. Compared to existing methods, this data-driven, template-free solution is more robust to variations in acquisition protocol, scanner vendor, and lesion burden. It represents a substantial advance in the search for clinically robust CT-based WMH segmentation.

### 4.3 AI Model Explainability

By evaluating the differential effects of imaging modality, time elapsed between the scans that provided the reference segmentations and the scans where the data were segmented, size and subtype of lesions with similar appearances but different aetiology, and overall WMH burden on segmentation performance we inform on the main source of spatial and volumetric disagreements between reference and resultant WMH masks, therefore explaining the model’s behaviour.

We found that modality-related differences, rather than temporal factors are the main drivers of variations in the accuracy of WMH volume estimates. The CT-MRI versus MRI-MRI group comparisons, supported by non-parametric testing and multivariable regression analyses, revealed that CT acquisitions at V0 produced systematically higher WMH volume estimates compared to the reference segmentations at V1 than those imaged with MRI at V0. The recognition of these modality-specific influences is critical to accurately assess the longitudinal burden of WMH and to guide methodological standardisation in large-scale multi-modal longitudinal imaging studies.

The proposed WMH segmentation model performs reasonably well in the presence of stroke lesions but large stroke lesions in the acute phase are a specific source of segmentation difficulty. This highlights the need to consider underlying cerebrovascular pathology while segmenting WMH. Future work may mitigate these challenges by using specialised training strategies or multimodal data integration to improve the reliability of automated WMH assessment in complex neurological contexts. Another approach could be to consider the WMH volume in the contralateral hemisphere as a proxy for the WMH volume in the ipsilateral hemisphere, to avoid ischaemic stroke lesions that may coalesce with the WMH influence on the measurements.

Beyond modality and stroke-related effects, our analyses suggest that PVS burden is associated with region-specific false-positive WMH behaviour. PVS are fluid-filled spaces, crucial for draining metabolic waste from the brain, that in many cases are topologically related with WMH [32]. Vascular stiffening and altered pulsatility, both of which alter fluid transport and are associated with WMH [36], have been thought to be key drivers of PVS enlargement. Moreover, blood-brain-barrier impairment, protein aggregation, neuroinflammation, and oxidative stress, also associated with WMH [37–39], all contribute to disrupt fluid dynamics and could potentially cause PVS enlargement [40]. Although individual PVS are typically assessed on MRI and are not reliably resolved on routine non-contrast CT, when they aggregate, interconnect, and are abundant, they may mimic WMH or confound WMH assessment. Consistent with this hypothesis, higher PVS density was associated with increased false-positive burden within both BG and CSO ROI; Figure 14). Moreover, the fraction of whole-volume false positives occurring within BG/CSO increased with regional PVS density. These findings motivate PVS-aware training and evaluation strategies (e.g., ROI-targeted hard-negative mining, region-specific calibration, or multi-task learning when PVS annotations are available) to reduce systematic false positives in BG/CSO while preserving sensitivity to true WMH.

### 4.4 Clinical Relevance

A robust CT-based WMH segmentation framework is advantageous for neuroradiological support systems. CT is often the first-line neuroimaging modality in many acute and emergency settings [7]. CT assessment of WMH could enhance risk stratification, facilitate rapid therapeutic decision-making, and direct patient management when MRI is contraindicated or unavailable. Additionally, automated WMH quantification on CT will facilitate large-scale studies and clinical trials with objective, reproducible measures of WMH burden. This will be important in evaluating the effects of treatment and better prognosis in patients with cerebrovascular diseases. Although the accuracy in delineating WMH volumes when these are small is still poor, the possibility of detecting the moderate to severe WMH in CT scans is still valuable for clinicians and patients.

In addition, the strong correlation between automated WMH burden estimates and Fazekas visual grades in MSS3 provides face-validity with established clinical WMH severity ratings, supporting the use of CT-derived WMH burden as a clinically interpretable quantitative marker when MRI is unavailable.

CT scans performed acutely at initial diagnosis are more susceptible to movement artefacts and suboptimal head positioning, given the clinical urgency inherent to acute presentations. This represents a systematic limitation affecting CT more than MRI in such settings. If computational analysis tools are to be applied in clinical practice, they will need to deal with poor-quality scan cases.

Our findings methodologically underscore that model training and validation protocols should consider modality-specific characteristics. We also found that large or recent stroke lesions can complicate WMH delineation. Domain adaptation [41] or multi-task learning approaches may be included in future modelling strategies to distinguish WMHs from other pathology-related signal changes and enhance model performance in complex clinical settings.

### 4.5 Limitations and Future Works

These advances are not without challenges. The delineation of mild and small WMH continue to be a difficult task because inherent low tissue contrast in CT images often blurs subtle differences into noise. Improvements in model performance could be further achieved by increasing the number or quality of mild WMH samples or by using image enhancement techniques or semi-supervised learning.

In addition, the intrinsic nature of CT image acquisition yields 3D volumes with non-uniform slice thickness, which is incompatible with the nifti-1 format used for processing. Further research into the development of an image format that is more inclusive and eliminates the distortions introduced by resampling the originally-acquired slices, which cannot be overcome at present by applying non-linear warping techniques, is necessary.

A third limitation is the reliance on pseudo-labelled data, which ultimately introduces a level of uncertainty in the reference data transferred into the CT model. This issue can be mitigated using iterative label refinement, e.g. via active learning strategies.

Moreover, the generality of the model has to be confirmed by validating the model performance on independent cohorts. Harmonising data across different scanning protocols may also improve robustness, e.g., by intensity normalisation or style transfer methods. Finally, integrating clinical covariates (e.g., vascular risk factors or genetic predispositions) with multi-modal imaging data may help us to better understand the pathophysiology and prognostic potential of WMH.

Future work should evaluate whether explicit PVS-aware modelling or post-processing reduces BG/CSO false positives without compromising sensitivity to true WMH.

## Data Availability

The image data used in this manuscript can be accessed only for research purposes by request to the principal investigators of the primary studies that provided the data. The framework presented uses software modules and libraries publicly available. The backbone neural network architecture used, namely nnUNet, is also freely available. The neural network weights of the proposed model can also be made available upon request to the authors.

https://github.com/rordenlab/dcm2niix

https://surfer.nmr.mgh.harvard.edu/docs/synthstrip/

https://cmictig.cs.ucl.ac.uk/wiki/index.php/NiftyReg

https://surfer.nmr.mgh.harvard.edu/

https://github.com/MIC-DKFZ/nnUNet

https://github.com/bjin96/ct-processing

## Data and Code Availability

The framework presented uses software modules and libraries publicly available. The backbone neural network architecture used, namely nnUNet, is also freely available. The imaging data could be available upon request to the principal investigators of the primary studies. The neural network weights of the proposed model can also be made available upon request.

## CRediT authorship contribution statement

**Nada Alamoudi:** Writing – Original Draft, Investigation, Methodology, Software, Formal Analysis, Data Curation, Validation, Visualisation, Conceptualisation, Funding Acquisition. **Maria Del C. Valdés Hernández:** Writing – Review & Editing, Supervision, Resources, Methodology, Formal Analysis, Data Curation, Validation, Visualisation, Conceptualisation, Project Administration. **Sohan Seth:** Writing – Review & Editing, Supervision, Conceptualisation. **Benjamin Jin:** Investigation and Visualisation (analysis/illustration of DICOM-to-NIfTI resampling effects; Section 2.3.2 and Supplementary Figure 1), Writing – Review & Editing. **Eleni Sakka; Carmen Arteaga-Reyes; Daniela Jaime García; Yajun Cheng; Angela C.C. Jochems:** Writing – Review & Approval, Data Curation. **Grant Mair:** Writing – Review & Editing. **Joanna M. Wardlaw:** Writing – Review & Editing, Data Curation, Resources. **Miguel O. Bernabeu:** Writing – Review & Editing, Supervision, Resources, Methodology, Validation, Conceptualisation.

## Acknowledgements and Funding

We thank the patients and volunteers who provided the images to the primary studies that contributed data to this study.

We also thank the large IST-3 Collaborative group (156 hospitals in 12 countries) - full details and primary results of the trial were published in 2012 [20], the MSS3 Study Group [19], the research radiographers and, in particular: Dr. Una Clancy for her work in the recruitment for the MSS3, Ms. Agniete Kampaite for the manual delineations of WMH in the MSS3, Dr. Stewart Wiseman for manually delineating the stroke lesions for the MSS3, and Drs. Michael J Thrippleton and Michael Stringer for their work setting-up the imaging protocol in the MSS3.

Nada Alamoudi gratefully acknowledges financial support from Taibah University, Kingdom of Saudi Arabia, under the PhD scholarship program. The IST-3 study was co-sponsored by The University of Edinburgh and the Lothian Health Board. The start-up phase was supported by a grant from the Stroke Association, UK. The expansion phase was funded by The Health Foundation UK. It also received funds from the following organisations: UK MRC (grant numbers G0400069 and EME 09-800-15) and managed by NIHR on behalf of the MRC-NIHR partnership; The Research Council of Norway; AFA Insurances (Sweden); the Swedish Heart Lung Fund; The Foundation of Marianne and Marcus Wallenberg; Stockholm County Council and Karolinska Institute Joint ALF-project grants (Sweden); the Government of Poland (grant number 2PO5B10928); the Australian Heart Foundation (grant number G 04S 1638); Australian NHMRC (grant number 457343); the Swiss National Research Foundation; the Swiss Heart Foundation; Foundation for health and cardio-/neurovascular research, Basel, Switzerland; the Assessorato alla Sanita, Regione dell’Umbria; Danube University, Krems, Austria. The imaging work was undertaken at the Brain Imaging Research Centre, a member of the SINAPSE collaboration, at the Division of Clinical Neurosciences, University of Edinburgh. SINAPSE is funded by the Scottish Funding Council and the Chief Scientist Offi ce of the Scottish Executive. Additional support was received from Chest Heart and Stroke Scotland, DesAcc, University of Edinburgh, Danderyd Hospital R&D Department, Karolinska Institutet, Oslo University Hospital, and the Dalhousie University Internal Medicine Research Fund.

The MSS3 study is funded by the UK Dementia Research Institute, which receives its funding from DRI Ltd, funded by the UK MRC, Alzheimer’s Society and Alzheimer’s Research UK; the Fondation Leducq Network for the Study of Perivascular Spaces in Small Vessel Disease (16 CVD 05); Stroke Association ‘Small Vessel Disease Spotlight on Symptoms (SVD-SOS)’(SAPG 19/100068) and Post-Doctoral Fellowship (SAPDF 18/100026), the Mexican National Council of Humanities Sciences and Technology (CONAHCYT, 2021-000007-01EXTF-00234, CAR), and The Row Fogo Charitable Trust Centre for Research into Aging and the Brain (BRO-D.FID3668413).

